# Demographic Characteristics, BAV Sievers Types, and Interactions Between BAV-Associated Valvulopathy and Aortopathy Differently Affect BAV Comorbidities and Outcomes of Aortic Valve Replacement

**DOI:** 10.1101/2020.08.12.20172171

**Authors:** Yijia Li, Qiong Zhao, Yue Qi, Yichen Qu, Akshay Kumar, Yan Yang, Xiongwen Chen

## Abstract

**Background:** Bicuspid aortic valve (BAV) is a common congenital disorder. The relationship between demographic and clinical characteristics, BAV Sievers types, BAV associated valvulopathy and/or aortopathy and outcomes of aortic valve replacement (AVR) are interwoven and complicate and have not been fully elucidated. We sought to find these interactions in a large cohort of BAV patients.

**Methods:** We retrospectively reviewed the data of 992 BAV patients and collected the complete demographic and clinical data (baseline characteristic, BAV Sievers types, BAV valvulopathy and aortopathy, and pre-, intra- and postoperative data) to comprehensively analyze these relationships.

**Results:** In 992 BAV patients, sex differences could be found in demography (body surface area [BSA], age and serum triglyceride), comorbidities, cardiac performance (left ventricular dimension and ejection fraction,), valvulopathy and aortopathy. Sievers types had the same distribution among male and female patients, and had an impact on the incidence of valvulopathy and aortopathy. In the entire cohort, the factors associated with valvulopathy included age, sex, BSA, systolic blood pressure (SBP) and aortopathy, while factors associated with aortopathy were age, sex, BSA and valvulopathy. Aortopathy and valvulopathy promoted the occurrence of each other. Similar risk factors for valvulopathy and aortopathy in male patients were found. For 658 BAV patients underwent AVR, the preoperative demographic characteristics were similar to the whole cohort. More males were required to have simultaneous ascending aortic replacement (AAR). For postoperative early adverse events (EAE) and total ICU hours > 24 hours, the only predict factors were age and aortic cross clamp (ACC) time, while LVEF changes (including postoperative LVEF <50%, LVEF increase or decrease more or less than 5% or 10%) were related to sex, SBP, preoperative LVEF, valvulopathy and aortopathy, AAR, ACC time. Postoperative length of stay > 7 days could be affected by SBP, AAR, aortic stenosis and ACC time.

**Conclusion:** Our study revealed comprehensive relationships between demographic characteristics, BAV Sievers types, valvulopathy and aortopathy, and the possible risk factors for adverse outcomes after AVR in BAV patients. Sex, SBP, age, Sievers types, subtypes and interactions between aortopathy and valvulopathy differently impact on aortopathy, valvulopathy and the short outcomes of AVR.

## Introduction

Bicuspid aortic valve (BAV) is a common congenital disorder with a prevalence rate of 2% in the general population, with male predominance. These patients have two aortic valves instead of three as in normal people. While BAV has little impact on the lives of young patients, over time it may cause severe cardiovascular complications such as aortic valve stenosis (AS), aortic valve regurgitation (AR) and aorta dilation. Those BAV comorbidities may require surgical interventions of the valves, such as aortic valve replacement (AVR), and/or the aorta, ^1, 2^.

It has been reported BAV Sievers types are associated with some demographic characteristics such as the sex. Krepp et al. reported that type 1 BAV with fusion between the left and right coronary cusp (type 1 L - R) was found more frequently in male than in female patients, whereas type 1 BAV with fusion between the left and noncoronary (type L - N) cusp was more common in female patients^3^. However, studies by others did not showed this difference^4, 5^. Demographic characteristics and Sievers types also have effects on BAV valvulopathy and aortopathy, which are the common complications in BAV patients. For the valvulopathy, male and female BAV patients have higher incidence AR and AS, respectively^4, 6^. Age was another independent determinant for BAV valvulopathy. Male BAV patients more often had moderate/severe AR at a young age, and the occurrence of AR decreased with age, while female BAV patients with had higher incidence of AS regardless of age^7^. Valve function was also affected by blood pressure and BAV Sievers type^5^. It becomes more complicated when considering BAV aortopathy and valvulopathy affects each other.

As for BAV aortopathy, age, male and BSA could be independent predictors of ascending aorta diameter in BAV patients^8^. Male patients have been reported to more frequently have isolated dilation of the aortic root and diffuse dilation of the aorta when compared with female patients^4, 6^. Higher BSA, female, age and AS are related to ascending aortic dilation^9^. Male patients with type 1 L - R are associated with larger sinus of Valsalva (SOV) diameter and AR, while female patients have a higher prevalence of AS^3^.

Furthermore, studies showed inconsistent risk factors for outcomes in BAV patients underwent AVR. One study implied that both short- and long-term outcomes were not significantly different between males and females^6^, while other suggested that females exhibit a higher relative risk of death, higher operative risk and longer lengths of hospital stay^10^. Valvulopathy also affects the postoperative left ventricular ejection fraction (LVEF) as the BAV-AS patients had a significant increase in LVEF when compared with BAV-AR patients after AVR^11^. However, it is unclear what are the other factors affecting the improvement or deterioration of LVEF after AVR.

To date, comprehensive analyses of the relationships between demographic (e.g., age, sex, body surface area (BSA)) and clinical (e.g., hypertension, diabetic mellites) characteristics, BAV Sievers types, incidences of different subtypes valvulopathy and/or aortopathy and their interactions, and the outcomes of aortic valve replacement (AVR) have not been done. In this study, we retrospectively reviewed a large number of BAV patients from January 2008 to December 2017 from our institution to elucidate these relationships.

## Methods

The Ethics Committees of Beijing Anzhen Hospital, Capital Medical University approved this retrospective study (2017028X) and waived the requirement for written patient informed consent form.

### Study Population and protocol

From January 2008 to December 2017, 1166 hospitalized patients from all departments of Beijing Anzhen Hospital with an echocardiographic diagnosis of BAV were screened and complete clinical record data were retrospectively reviewed and collected. Nine hundred and ninety-two BAV patients were enrolled in our study after excluding patients (n = 174) with preoperative aortic dissection, combined congenital heart disease and/or connective tissue disorders, including Marfan syndrome and Ehlers-Danlos syndrome. Among these patients, 658 BAV patients underwent AVR. We first analyzed the relationships between demographic characteristics, BAV Sievers types, overall and different subtypes aortopathy, overall and different subtypes valvulopathy, and short-term post-operative outcomes (Figure 1).

**Figure 1.**
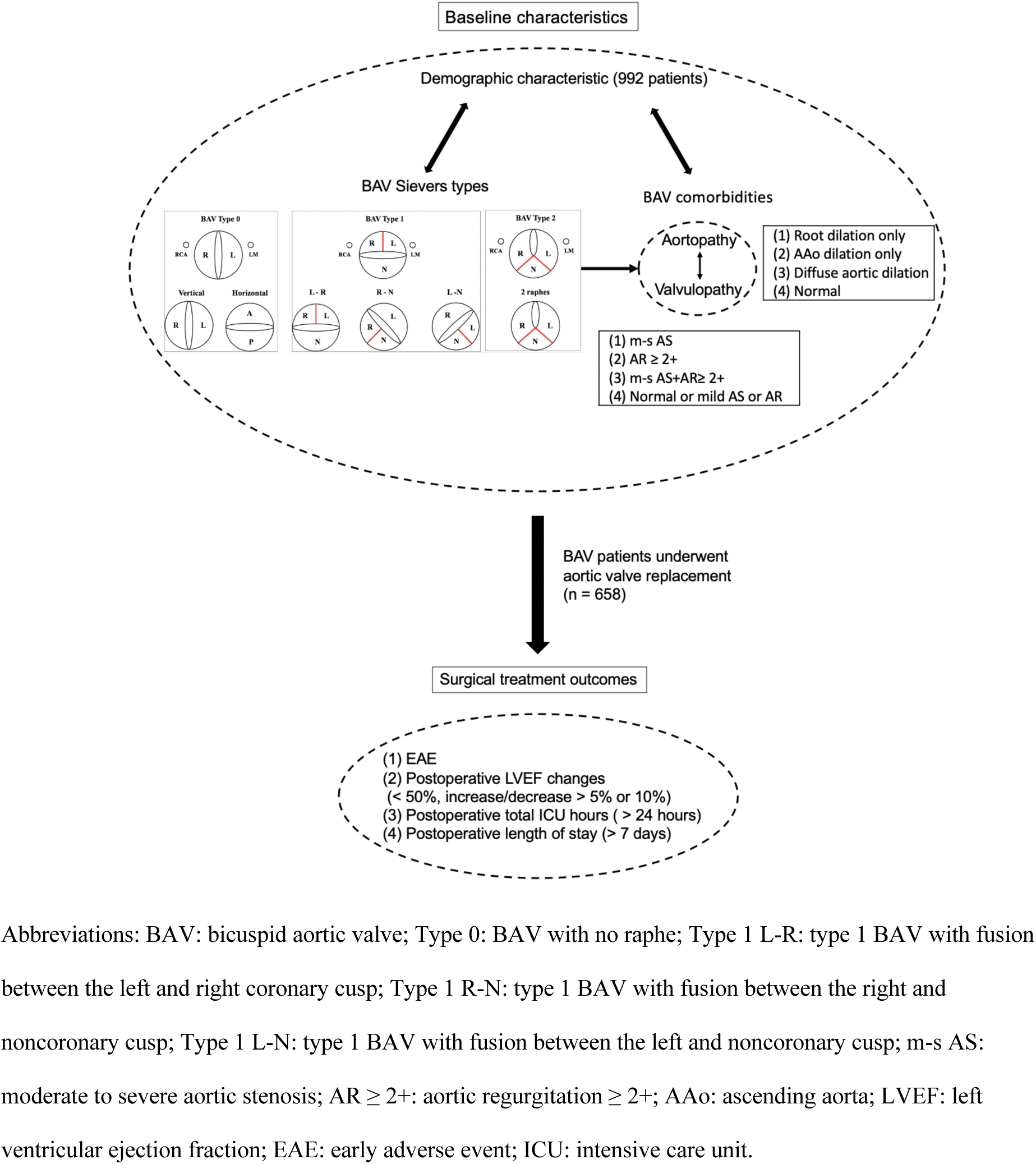
Study Design.

### Data Collection

Complete clinical record data including demographics, clinical history, medical history, physical examination, imaging examination, laboratory data, and clinical treatments were queried retrospectively. Hypertension was considered as a diagnosis when it was newly diagnosed or treated with medicine within 6 months before echocardiography examination.

For patients underwent AVR, additional information including preoperative drug use, intra- and postoperative parameters, like postoperative early adverse event (EAE), postoperative LVEF, total intensive care unit (ICU) hours and postoperative length of stay were recorded. EAE encompasses in-hospital and 30-day mortality, major cardiac events (myocardial infarction, ventricular fibrillation, acute heart failure, use of circulatory support like intra-aortic balloon pump [IABP] or extracorporeal membrane oxygenation [ECMO]), acute renal failure, stroke and reoperation for bleeding).

### Echocardiography Data

Echocardiography measurements were performed according to the guidelines of American Society of Echocardiography (ASE), European Society of cardiology (ESC) and American College of Cardiology/American Heart Association (ACC/AHA)^12-14^. All patients went through comprehensive transthoracic echocardiography evaluation, and those with the diagnosis of BAV by echocardiography were included for this study. BAV was diagnosed by parasternal long- and short-axis view, showing the existence of only 2 commissures in systole, and was classified based on the Sievers classification system (Sievers types)^15^ as type 0 (BAV with no raphe), type 1 L-R (BAV with fusion between the left and right coronary cusp), type 1 R-N (BAV with fusion between the right and noncoronary cusp), type 1 L-N (BAV with fusion between the left and noncoronary cusp). There was no type 2 BAV patient in our cohort.

Valvular dysfunction (valvulopathy, including AS and/or AR) was determined with hemodynamic measurements and the degrees of AS and AR were scored based on the ESC and ACC/AHA guidelines for the management of valvular heart disease^13, 14, 16^. Valvular function was categorized into four conditions: (1) moderate to severe aortic valve stenosis (m–s AS) only, (2) aortic valve regurgitation ≥ 2+ (AR ≥ 2+) only, (3) m–s AS + AR ≥ 2+, (4) mild AS and/or mild AR or normal ^17^

For the evaluation of aortopathy, SOV and ascending aorta (AAo) were measured using the leading edge to leading edge technique in the parasternal long axis view perpendicular to the centerline of the aorta. BSA indexed SOV and AAo dimensions were calculated. The dilation of SOV or AAo was defined as the diameter ≥ 40mm according to the guidelines^18, 19^ and literature^20^.The classification of BAV aortopathy as follow: (1) aortic root (comprising of the SOV, aortic valve and coronary ostia) dilation only, (2) AAo dilation only, (3) diffuse aortic dilation (i.e., dilation of both aortic root and AAo) and (4) no dilation (normal)^21^.

### Statistical Analysis

Continuous variables, expressed as means ± standard deviation (SD) in case of normally distributed variables or as medians (interquartile ranges (IQR), i.e., median (Q1, Q3), were compared using unpaired Student’s *t* test or *Mann-Whitney U* test or one-way analysis of variance where appropriate. Categorical variables, expressed as numbers and/or percentages, were compared using the Chi-square or *Fisher’s* exact tests.

The hazard ratios (HR) of age, sex, valvulopathy and aortopathy and other parameters for postoperative EAE were calculated using proportional Cox hazard models. The odds ratios (OR) of basic clinical parameters, like age, sex, BSA, and BAV Sievers type, BAV-induced comorbidities (including valvulopathy and aortopathy), were analyzed with multivariable logistic regression models adjusted for significant covariables in univariate analysis with backward stepwise method, to investigate factors affecting valvulopathy, aortopathy and short-term post-operative outcomes. Statistical analyses were performed using SPSS version 25.0 (IBM Corp., Armonk, N.Y., USA). All tests were 2-sided and a *P* value of < 0.05 was considered statistically significant.

## Results

### Sex differences in demography, comorbidities, cardiac performance, Sievers types, valvulopathy and aortopathy in BAV patients

We enrolled 992 patients according to our inclusion criteria, in which 734 (74.0%) patients were male, and 258 (26.0%) patients were female, agreeing with BAV’s predominance in males^4^. Females were about 3.6 years older than males when first diagnosed with BAV in our institution. Weight, height and BSA were greater in males. The prevalence of tobacco use was higher in male patients. Male BAV patients had higher incidence of hypertension. Serum triglyceride was higher in male patients, but female patients had higher levels of total cholesterol, high-density lipoprotein (HDL) and low-density lipoprotein (LDL). LVEDD and LVESD were significantly smaller in female (LVEDD, 57.54 ± 11.90 mm vs. 47.67 ± 6.92 mm, *P* < 0.001; LVESD, 39.05 ± 10.62 mm vs. 30.88 ± 6.61 mm, *P* < 0.001) while LVEF was lower in males (59.79 ± 10.05 % vs. 63.74 ± 8.67 %, *P* < 0.001) (Table 1).

**Table 1.**
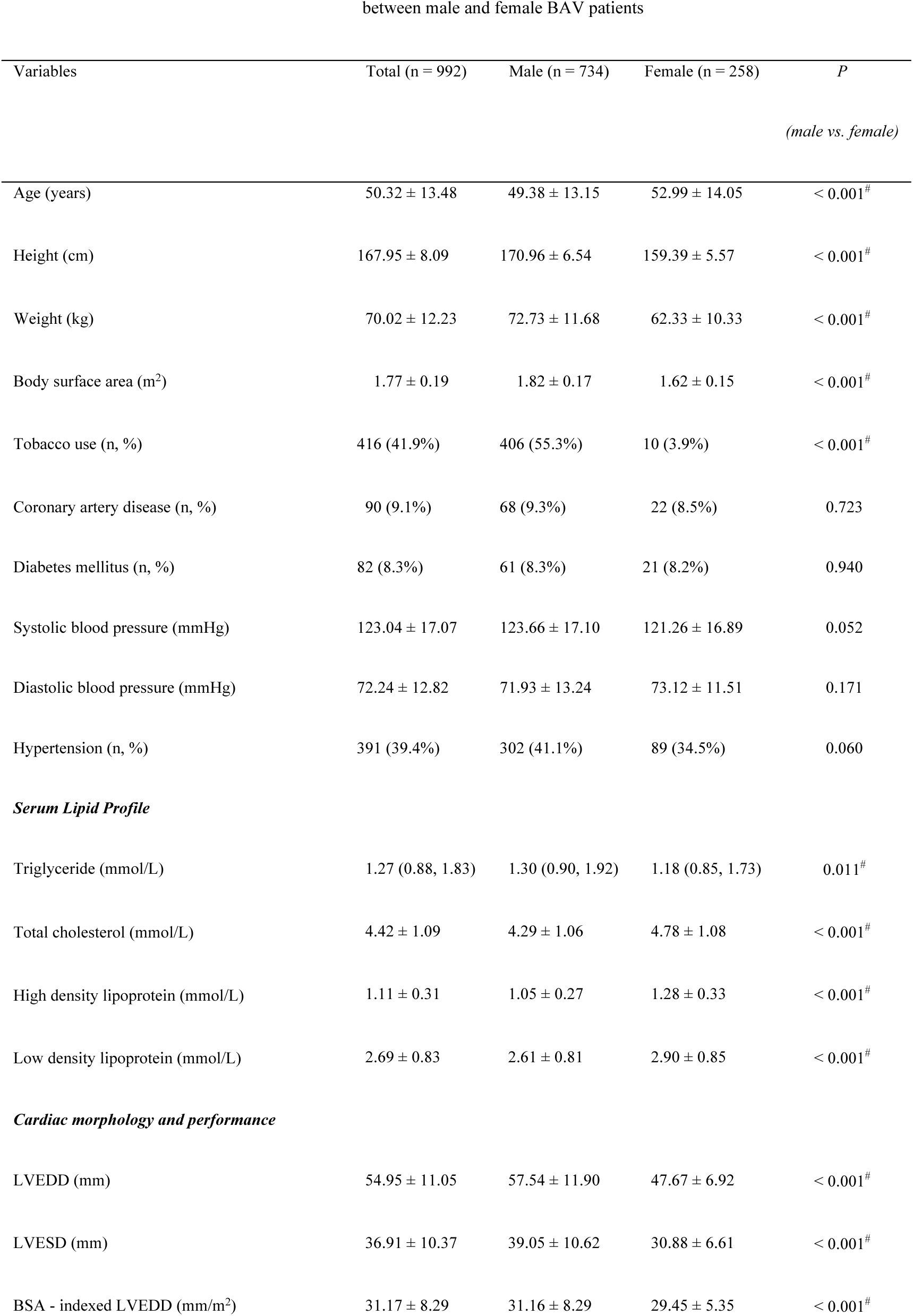
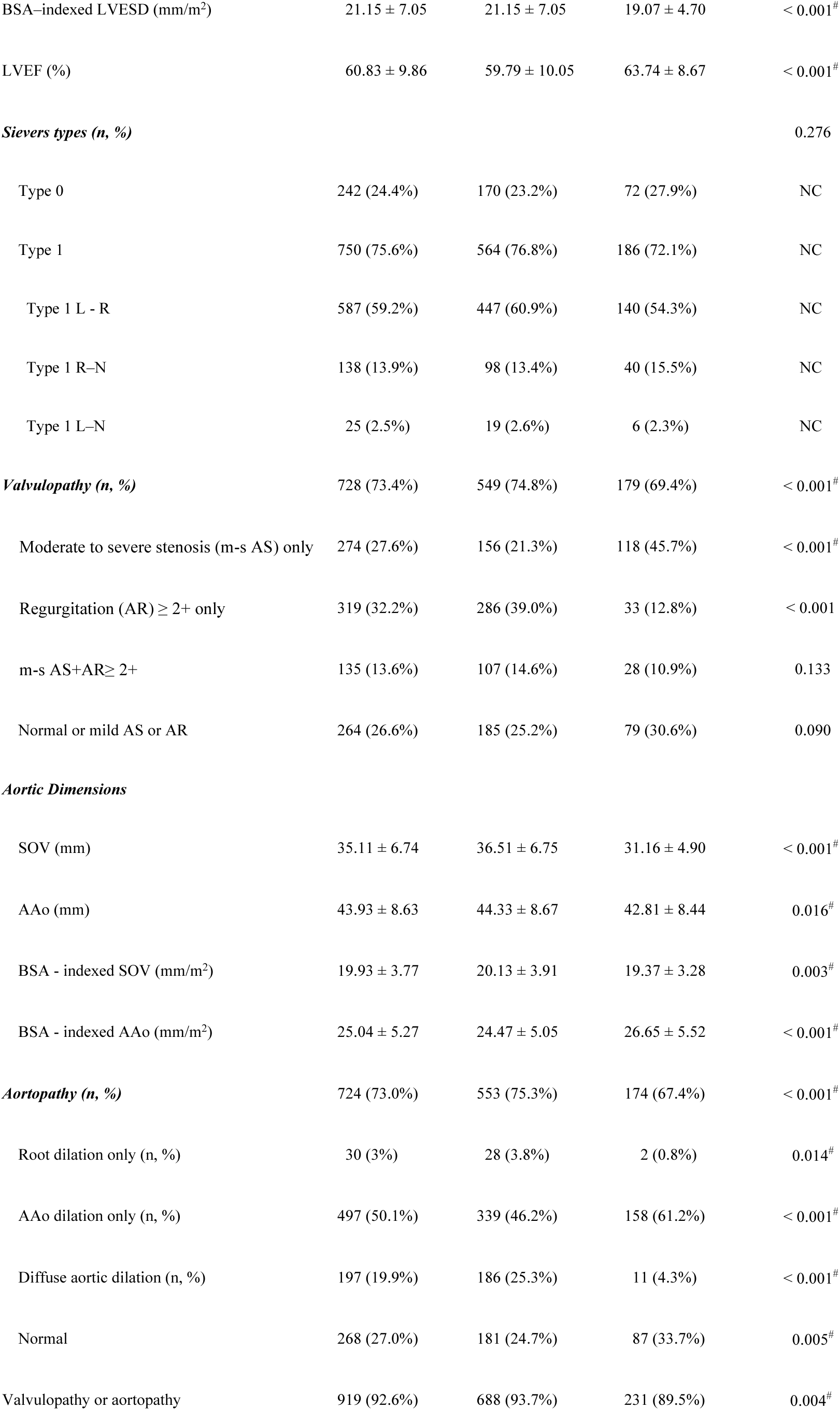
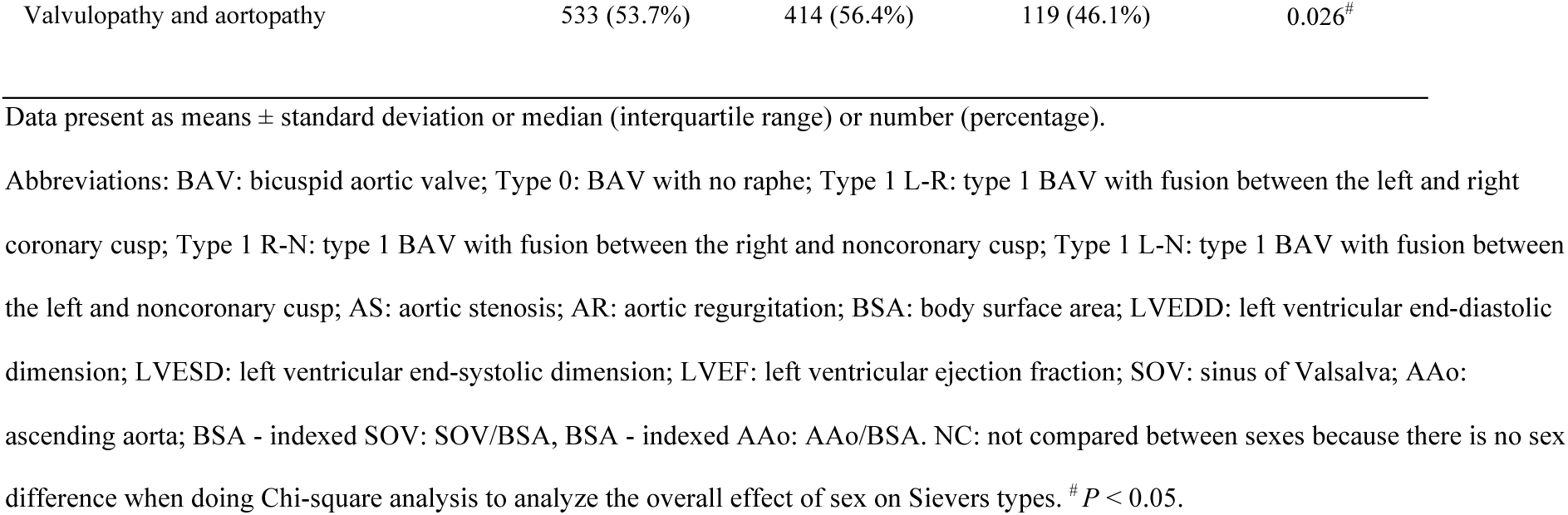
Differences in demography, clinical characteristics, Sievers types, valvulopathy and aortopathy between male and female BAV

In all 992 patients, the most common BAV Sievers types were type 1 L - R (59.2%), followed by type 0 (24.4%), type 1 R - N (13.9%), and type 1 L - N (2.5%). There was no sex difference in the distribution of Sievers types (Table 1 and Table 2).

**Table 2.** Demography and Clinical Characteristics in BAV patients with different Sievers types

| Variables | Type 0 (n = 242) | Type 1 (n = 750) |  |  | *P*<br>(different Sievers types) |
| --- | --- | --- | --- | --- | --- |
|  |  | Type 1 L-R (n = 587) | Type 1 R-N (n = 138) | Type 1 L-N (n = 25) |  |
| Age (years) | 49.85 ± 13.60 | 50.60 ± 13.52 | 49.75 ± 13.33 | 51.44 ± 12.53 | 0.759 |
| Male (n, %) | 170 (70.2%) | 447 (76.1%) | 98 (71.0%) | 19 (76.0%) | 0.276 |
| Height (cm) | 167.29 ± 8.46 | 168.38 ± 7.62 | 167.04 ± 9.38 | 168.92 ± 7.15 | 0.503 |
| Weight (kg) | 69.75 ± 12.86 | 70.25 ± 11.79 | 69.71 ± 12.60 | 68.92 ± 14.59 | 0.912 |
| Body surface area (m<sup>2</sup>) | 1.76 ± 0.20 | 1.77 ± 0.18 | 1.76 ± 0.20 | 1.76 ± 0.22 | 0.862 |
| Tobacco use (n, %) | 103 (42.6%) | 248 (42.2%) | 52 (37.7%) | 13 (52.0%) | 0.546 |
| Coronary artery disease (n, %) | 28 (11.6%) | 47 (8.0%) | 13 (9.4%) | 2 (8.0%) | 0.551 |
| Diabetes mellitus (n, %) | 23 (9.5%) | 45 (7.7%) | 13 (9.4%) | 1 (4.0%) | 0.654 |
| Systolic blood pressure (mmHg) | 122.50 ± 17.62 | 123.94 ± 16.70 | 122.41 ± 17.78 | 110.60 ± 10.87 | < 0.001<sup>#</sup> |
| Diastolic blood pressure (mmHg) | 75.15 ± 12.78 | 71.33 ± 12.84 | 71.54 ± 12.45 | 69.40 ± 10.65 | 0.001<sup>#</sup> |
| Hypertension (n, %) | 95 (39.3%) | 241 (41.1%) | 47 (34.1%) | 8 (32.0%) | 0.408 |
| **Serum Lipid Profile** |  |  |  |  |  |
| Triglyceride (mmol/L) | 1.22 (0.86, 1.77) | 1.27 (0.88, 1.88) | 1.30 (1.00, 1.76) | 1.32 (0.80, 2.30) | 0.682 |
| Total cholesterol (mmol/L) | 4.40 ± 1.02 | 4.43 ± 1.12 | 4.44 ± 1.08 | 4.20 ± 0.85 | 0.679 |
| High density lipoprotein (mmol/L) | 1.11 ± 0.32 | 1.11 ± 0.30 | 1.10 ± 0.34 | 1.03 ± 0.29 | 0.458 |
| Low density lipoprotein (mmol/L) | 2.67 ± 0.79 | 2.71 ± 0.84 | 2.67 ± 0.88 | 2.53 ± 0.72 | 0.770 |
| ***Cardiac morphology and performance*** |  |  |  |  |  |
| LVEDD (mm) | 53.17 ± 10.74 | 55.79 ± 11.39 | 55.08 ± 10.06 | 52.20 ± 9.16 | 0.004<sup>#</sup> |
| LVESD (mm) | 35.81 ± 10.38 | 37.58 ± 10.74 | 36.45 ± 8.89 | 34.82 ± 8.17 | 0.050 |
| BSA - indexed LVEDD (mm/m<sup>2</sup>) | 30.19 ± 7.22 | 30.81 ± 8.16 | 31.37 ± 6.49 | 29.86 ± 5.27 | 0.006<sup>#</sup> |
| BSA-indexed LVESD (mm/m<sup>2</sup>) | 20.35 ± 6.61 | 20.73 ± 6.91 | 20.66 ± 5.67 | 19.92 ± 4.84 | 0.109 |
| LVEF (%) | 60.90 ± 10.58 | 60.55 ± 10.00 | 61.91 ± 8.29 | 60.68 ± 6.97 | 0.621 |
| ***Valvulopathy (n, %)*** |  |  |  |  | < 0.001<sup>#</sup> |
| Moderate to severe stenosis (m-s AS) only | 70 (28.9%) | 166 (28.3%) | 32 (23.2%) | 6 (24.0%) | 0.603 |
| Regurgitation (AR) ≥ 2+ only | 47 (19.4%) | 219 (37.3%) | 48 (34.8%) | 5 (20.0%) | < 0.001<sup>#</sup> |
| m-s AS+AR ≥ 2+ | 24 (9.9%) | 81 (13.8%) | 26 (18.8%) | 4 (16.0%) | 0.104 |
| Normal or mild AS or AR | 101 (41.7%) | 121 (20.6%) | 32 (23.2%) | 10 (40.0%) | < 0.001<sup>#</sup> |
| ***Aortic Dimensions*** |  |  |  |  |  |
| SOV (mm) | 33.39 ± 6.46 | 36.11 ± 6.86 | 33.84 ± 6.06 | 35.51 ± 6.05 | < 0.001<sup>#</sup> |
| AAo (mm) | 44.30 ± 8.88 | 43.72 ± 8.77 | 44.10 ± 7.63 | 44.39 ± 8.64 | 0.221 |
| BSA - indexed SOV (mm/m<sup>2</sup>) | 18.97 ± 3.81 | 20.46 ± 3.74 | 19.30 ± 3.29 | 20.42 ± 4.17 | < 0.001<sup>#</sup> |
| BSA - indexed AAo (mm/m<sup>2</sup>) | 25.26 ± 5.53 | 24.85 ± 5.31 | 25.35 ± 4.60 | 25.45 ± 5.24 | 0.119 |
| ***Aortopathy (n, %)*** |  |  |  |  | 0.001<sup>#</sup> |
| Root dilation only (n, %) | 3 (1.2%) | 24 (4.1%) | 2 (1.4%) | 1 (4.0%) | 0.105 |
| AAo dilation only (n, %) | 139 (57.4%) | 262 (44.6%) | 84 (60.9%) | 12 (48.0%) | < 0.001<sup>#</sup> |
| Diffuse aortic dilation (n, %) | 33 (13.6%) | 139 (23.7%) | 20 (14.5%) | 5 (20%) | 0.003<sup>#</sup> |
| Normal | 67 (27.7%) | 162 (27.6%) | 32 (28.0%) | 7 (28.0%) | 0.755 |
| Valvulopathy or aortopathy | 209 (86.4%) | 554 (94.4%) | 113 (96.4%) | 23 (92.0%) | < 0.001<sup>#</sup> |
| Valvulopathy and aortopathy | 107 (44.2) | 337 (57.4%) | 79 (57.2%) | 10 (40.0%) | 0.002<sup>#</sup> |

Data present as means ± standard deviation or median (interquartile range) or number (percentage).
Abbreviations: BAV: bicuspid aortic valve; Type 0: BAV with no raphe; Type 1 L-R: type 1 BAV with fusion between the left and right coronary cusp; Type 1 R-N: type 1 BAV with fusion between the right and noncoronary cusp; Type 1 L-N: type 1 BAV with fusion between the left and noncoronary cusp; AS: aortic stenosis; AR: aortic regurgitation; BSA: body surface area; LVEDD: left ventricular end-diastolic dimension; LVESD: left ventricular end-systolic dimension; LVEF: left ventricular ejection fraction; SOV: sinus of Valsalva; AAo: ascending aorta; BSA - indexed SOV: SOV / BSA, BSA - indexed AAo: AAo / BSA. <sup>#</sup> $P < 0.05$ .

BAV strongly predisposes the patients to aortic and/or valvular diseases. The percentage of patients with at least of one form of valvulopathy and/or aortopathy in our cohort was 92.6% and 53.7% had both one form of aortopathy and one form of valvulopathy. Males had higher percentage of at least one form of valvulopathy or aortopathy, and higher percentage of combined valvulopathy and aortopathy (Table 1).

The overall prevalence of valvulopathy in all BAV patients was high (73.4%), among which 75.4% were males and 24.6% were females. Among all BAV patients, males were more frequently having AR ≥ 2+ only than females (39.0% vs. 12.8%, *P* < 0.001), whereas females were prone to have m-s AS only (45.7% vs 21.3%, *P* < 0.001) (Table 1).

As shown in Table 1, aortopathy was common in our cohort of BAV patients (73.0%) and most common aortopathy was AAo dilation only (50.1%), followed by diffuse aortic dilation (19.9%), and root dilation only (3.0%). Compared to females, male patients showed higher incidence of aortic diffuse dilation (25.3% vs. 4.3%, *P* < 0.001) and root dilation (3.8% vs. 0.8%, *P* = 0.014);, while female patients had greater incidence of AAo dilation only (61.2% vs. 46.2%, *P* < 0.001) (Table 1). The linear dimension of SOV, AAo, and BSA indexed SOV were significantly larger in males when compared with females (SOV, 36.51 ± 6.75mm vs. 31.16 ± 4.90mm, *P* < 0.001; SOV/BSA, 20.13 ± 3.91mm/m^2^ vs. 19.37 ± 3.28mm/ m^2^, *P* = 0.003; AAo, 44.33 ± 8.67mm vs. 42.81 ± 8.44mm, *P* = 0.016), but BSA indexed AAo was significantly larger in females instead (24.47 ± 5.05 vs. 26.65 ± 5.52, *P* < 0.001), which indicated that females had larger AAo diameter when shared the same BSA as males.

#### The association between different Sievers types with clinical characteristics, valvulopathy and aortopathy in BAV patients

Patients with different BAV Sievers types were equipped with distinct clinical characteristics including DBP, SBP, LVEDD, LVESD, and BSA normalized LVEDD or LVESD. Patients with type 0 Sievers type had the highest diastolic blood pressure (DBP), while type 1 L-N had lowest SBP and DBP. Type 1 L-R and type 1 R-N had larger LV diameter than type 0 and type L-N. Type 1 L-N also had the smallest LVEDD/BSA (Table 2).

In our cohorts, Sievers types had an impact on the incidence of valvulopathy and aortopathy. For valvulopathy, the incidence of AR ≥ 2+ was greater in type 1 L-R (37.3%) and type 1 R-N (34.8%) than in type 0 (19.4%) and type 1 R-N (20.0%). Interestingly, type 0 and type 1 R-N had most normal or mild AS or AR (Table 2). For BAV aortopathy, type 1 L-R and type 1 L-N had larger SOV diameter and BSA indexed SOV than type 0 and type 1 R-N. Type 0 and Type 1 R-N had higher frequency of AAo dilation only than type 1 L-R and type 1 L-N, which on the contrary had lower incidence of diffuse aortic dilation than type 0 and type 1 R-N (table 2). Type 1 L-R and type 1 R-N had higher percentage of at least one form of valvulopathy or aortopathy, and higher percentage of combined valvulopathy and aortopathy (Table 2).

#### Risk factors for valvulopathy in all, male and female BAV patients

We further identified independent risk factors for all these three types of valvulopathy, m-s AS only, AR ≥ 2+ only and both in all, male and female BAV patients, by adjusting for confounders. In the whole cohort, for m-s AS only, age predicted a risk while higher BSA and diffuse aortic dilation strongly decreased the risk of AS. For AR ≥ 2+ only, higher SBP (instead of hypertension history) and male increase its incidence, while type 1 L-R might be another potential risk factor (OR = 2.943, *P* = 0.066). As for m-s AS+AR ≥ 2+, greater BSA greatly decreased the risk, while AAo dilation only, diffuse aortic dilation and male were the risk factors (Table 3 and supplementary Table 1).

**Table 3.**
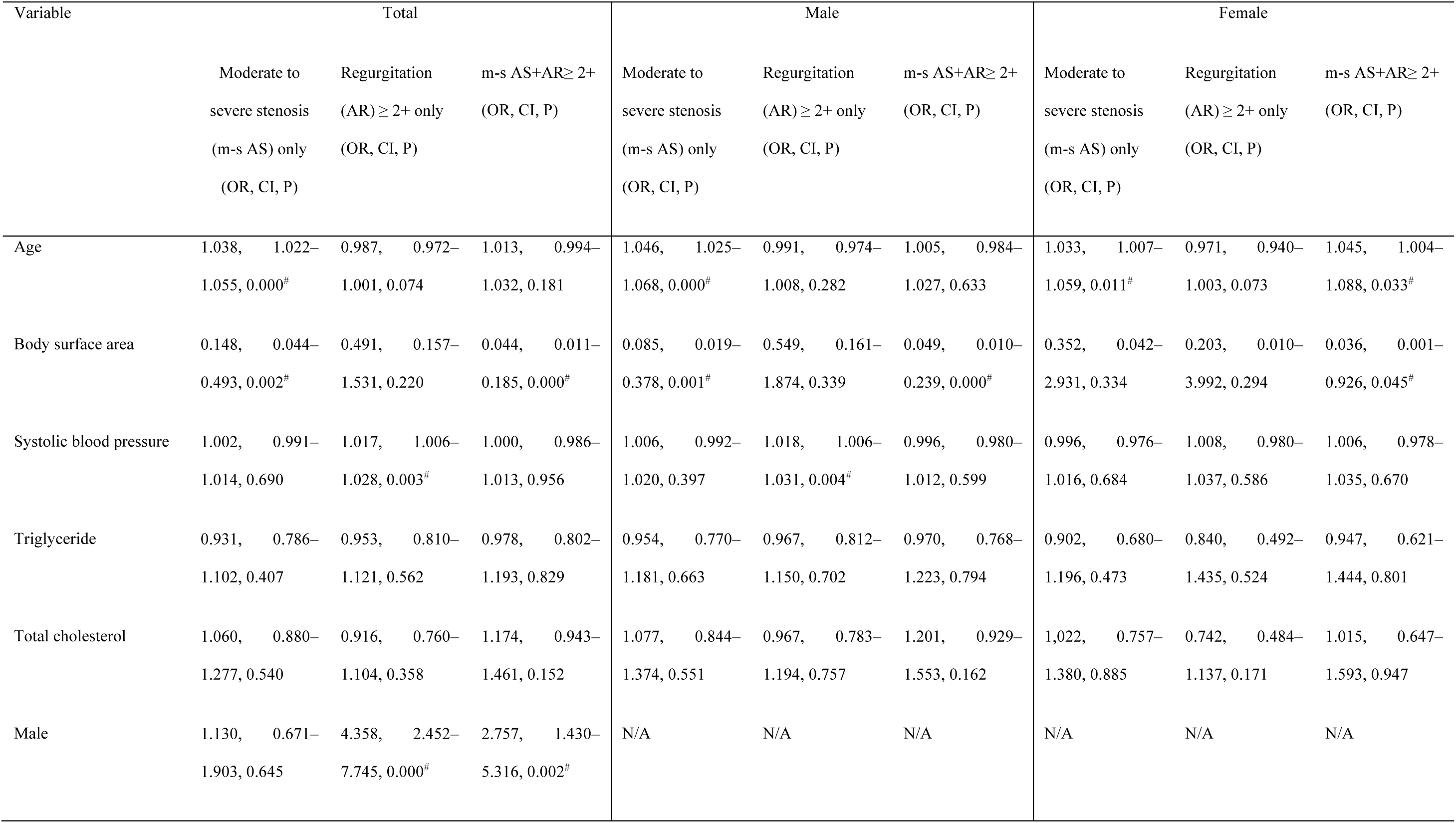
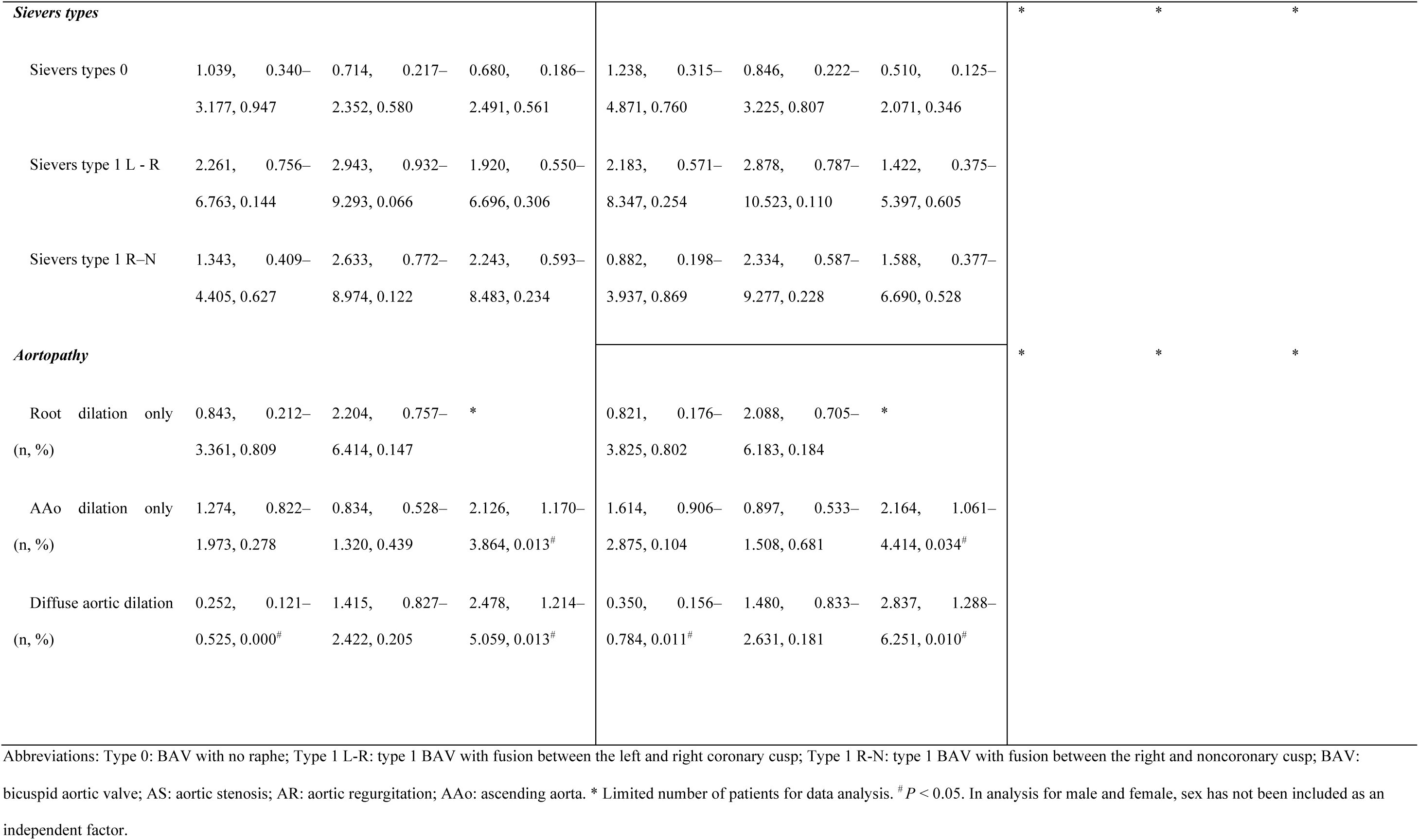
Risk factors for BAV Valvulopathy

We further looked into the risk and protective factors for BAV associated valvulopathy in male and female BAV patients. For male BAV patients, age was a predict factor (OR = 1.046, *P* < 0.001) for m-s AS only, while higher BSA (OR = 0.085, *P* = 0.001) decreased the risk of it. Higher SBP (OR = 1.018, *P* = 0.004) increased the incidence of AR ≥ 2+ only (Table 3 and supplementary Table 2). For m-s AS+AR ≥ 2+, higher BSA decreased the risk (OR = 0.049, *P* < 0.001).

There were interactions between valvulopathy and aortopathy in males: Males were prone to have diffuse aortic dilation (OR = 3.868, *P* = 0.001) (Table 4). Diffuse aortic dilation (OR = 0.350, *P* < 0.011) decreased the risk of m-s AS only; AAo dilation only (OR = 2.164, *P* = 0.034) and diffuse aortic dilation (OR = 2.837, *P* = 0.010) were the risk factors for m-s AS+AR ≥ 2+. In females, age is a predictor for m-s AS only (OR = 1.033, *P* = 0.011) and m-s AS + AR ≥ 2+ (OR = 1.045, *P* = 0.033), while higher BSA decrease the risk for m-s AS + AR ≥ 2+ (OR = 0.036, *P* = 0.045) (Table 3). For other factors, the numbers of affected female patients were too few and thus we could not assess the risk of them.

**Table 4.**
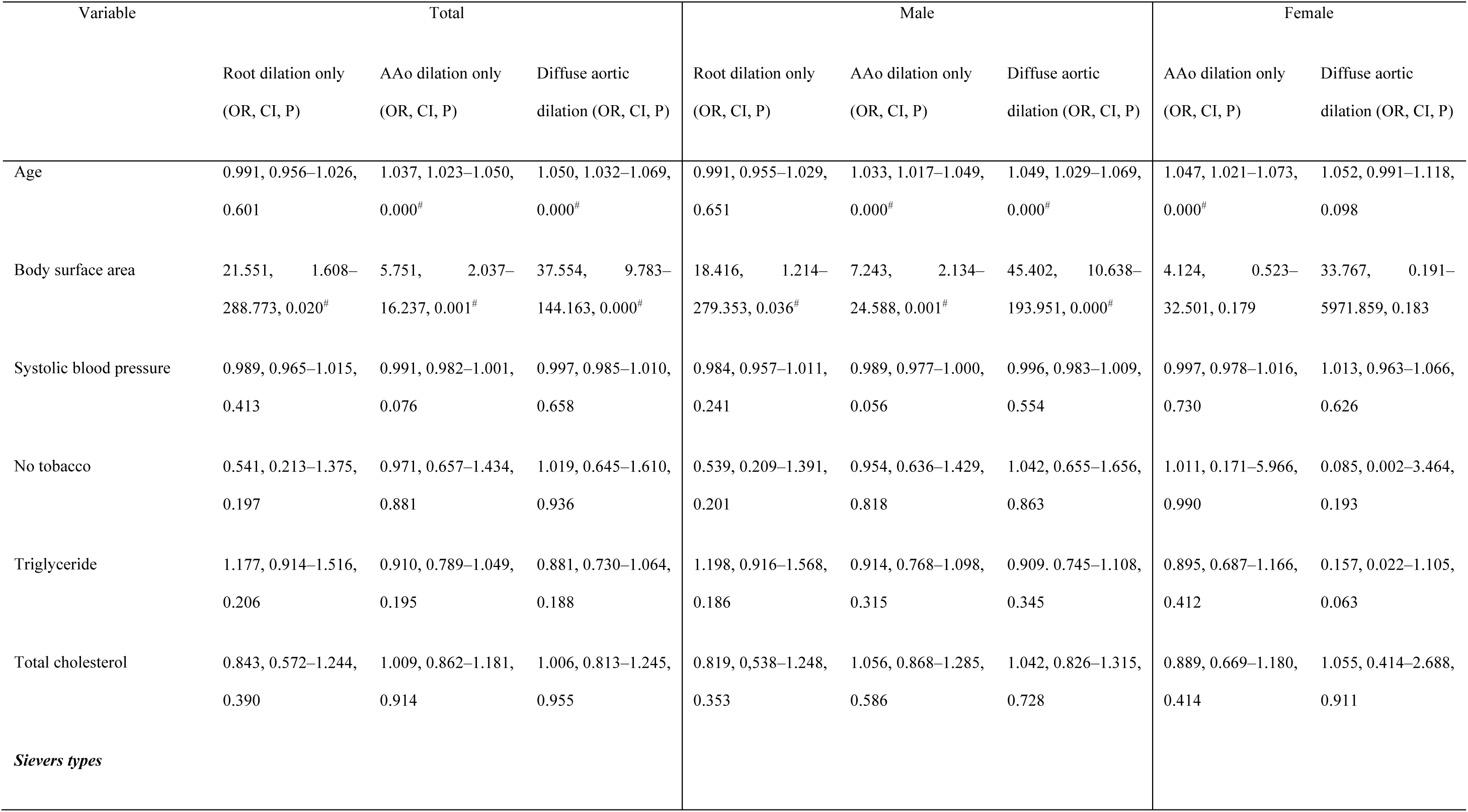
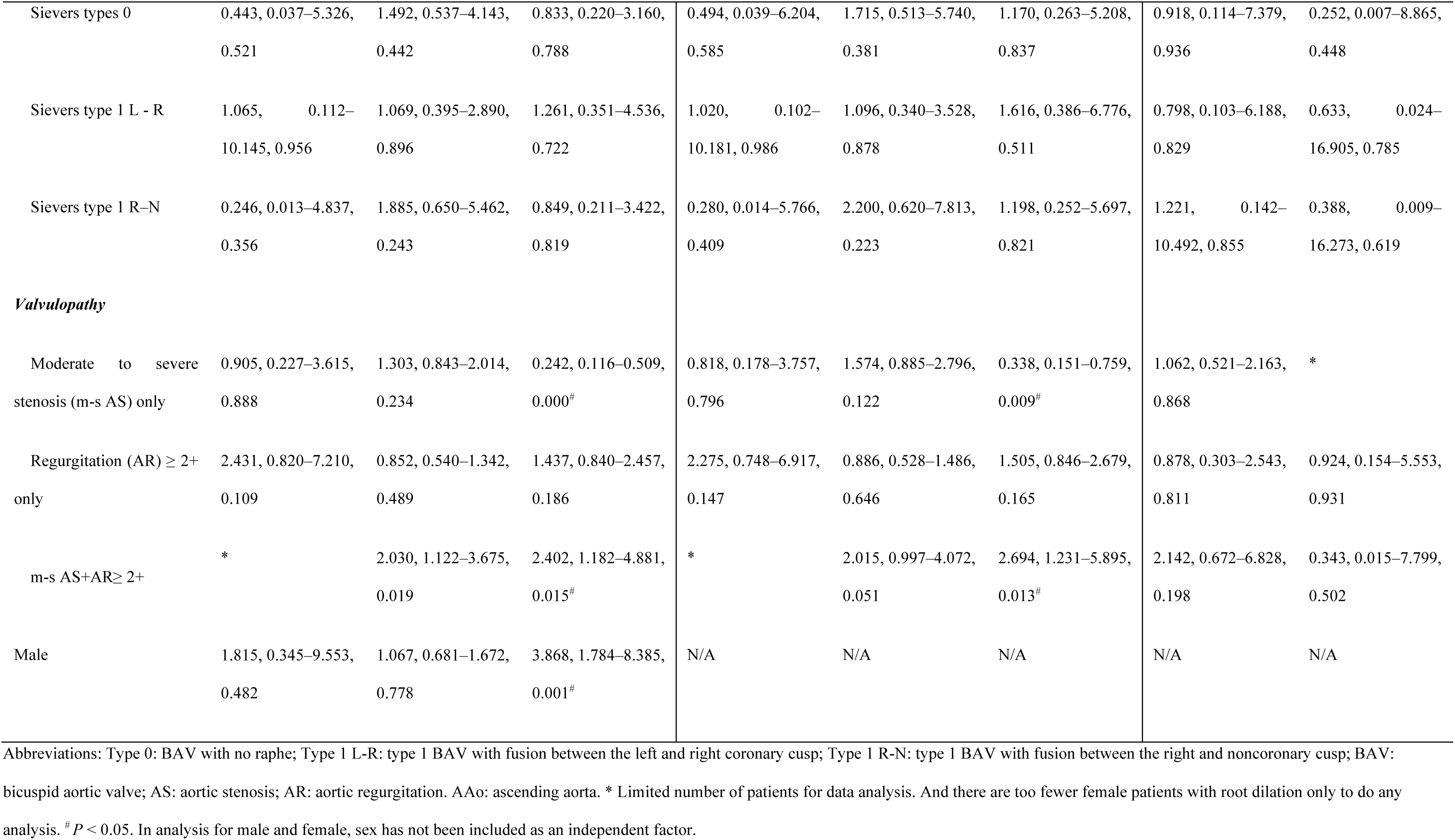
Risk factors for BAV aortopathy

### Risk factors for BAV aortopathy in all, male and female BAV patients

In the entire cohort, BSA was the only risk factor for all forms of aortopathy (aortic root dilation only, OR = 21.551, *P* = 0.020; AAo dilation only, OR = 5.751, *P* = 0.001; diffuse aortic dilation, OR = 37.554, *P* < 0.001). Age was a risk factor for AAo dilation only and diffuse aortic dilation in the whole cohort (AAo dilation only, OR = 1.037, *P* < 0.001; diffuse aortic dilation, OR = 1.050, *P* < 0.001) and male patients (AAo dilation only, OR = 1.033, *P* < 0.001; diffuse aortic dilation, OR = 1.049, *P* < 0.001). BSA was also the only risk factor for all three forms of aortopathy in males (aortic root dilation only, OR = 18.416, *P* = 0.036; AAo dilation only, OR = 7.243, *P* = 0.001; diffuse aortic dilation, OR = 45.402, *P* < 0.001), but not in females. In the whole cohort and males, valvulopathy affected the aortopathy: m-s AS only decreased the risk for diffuse aortic dilation in the whole study cohort (OR = 0.242, *P* < 0.001) and males (OR = 0.338, *P* < 0.001), while m-s AS + AR ≥ 2+ increase the risk for the whole cohort (OR = 2.402, *P* = 0.015) and males (OR = 2.694, *P* = 0.013). (Table 4). For females, age was a risk factor for AAo dilation only (OR = 1.047, *P* < 0.001). Because the numbers of female patients associated with some factors were limited, we could not perform additional analysis.

### Pre-, intra- and post-operative characteristics of all, male and female BAV patients underwent AVR

The preoperative demographic characteristics for BAV patients underwent AVR were similar as the whole cohort. In 658 BAV patients underwent AVR, 75.1% were males, while 24.9% were females. While most pre-operative demographic and clinical characteristics were similar between male and female patients, males were younger (48.74 ± 12.73 years vs. 55.34 ± 11. 20 years, *P* < 0.001) and had lower LVEF (60.67 ± 8.57 % vs. 63.67 ± 8.18 %, *P* < 0.001), suggesting more need of AVR in male patients. Males also had lower DBP (70.94 ± 13.35 mmHg vs. 73.30 ± 11.07 mmHg, *P* = 0.026), greater BSA (1.82 ± 0.16 m^2^ vs. 1.62 ± 0.15 m^2^, *P* < 0.001), higher frequency of AR ≥ 2+only (223 [45.1%] vs. 25 [15.2%], *P* < 0.001) and aortic root dilation only (19 [3.8%] vs. 1 [0.6%], *P* = 0.036) and diffuse aortic dilation (147 [29.8%] vs. 7 [4.3%], *P* < 0.001). Females were more likely to have m–s AS only (119 [24.1%] vs. 101 [61.6%], *P* < 0.001) and AAo dilation only (226 [45.7%] vs. 106 [64.6%], *P* < 0.001).

More male patients were required to have simultaneous ascending aortic replacement when they underwent AVR (248 [50.2%] vs. 62 [37.8%], *P* = 0.006), again suggesting more severe conditions in male BAV patients.

### Factors related to postoperative EAE, L VEF and L VEF changes after AVR

Age (HR = 1.030; *P* = 0.002) and longer aortic cross clamp (ACC) time (HR = 1.012, *P* < 0.001) were predict factors for EAE. Additionally, we used LVEF, total ICU hours, and postoperative hospital stay as indices of postoperative outcomes. It has been reported that post-operative LVEF<50% is associated adverse long-term prognosis^22^. Sex was a major factor affecting outcomes. Postoperatively, on average LVEF decreased in all, male and female patients compared to preoperative values and was lower in males than females (57.03 ± 10.11% vs. 60.83 ±10.66 %, *P* < 0.001). Female patients had longer total ICU hours (21 [19, 29.74] vs. 21 [18, 26], *P* = 0.042), and higher percentage of patients needing total ICU hours > 24 hours (38.4% vs. 29.8%, *P* = 0.039). Males had higher percentage of postoperative LVEF < 50% and lower percentage of postoperative LVEF increase > 10%. Multivariable logistic regression confirmed that male was the risk factor for postoperative LVEF < 50% (OR = 3.072, *P* = 0.033). Females stayed in hospital after surgery longer than males and had higher frequency of postoperative length of stay > 7 days. There were 13.7% patients showing EAE but the incidence of EAE was not differing significantly between sex (HR = 1.213, *P* = 0.441) (Table 5).

**Table 5.**
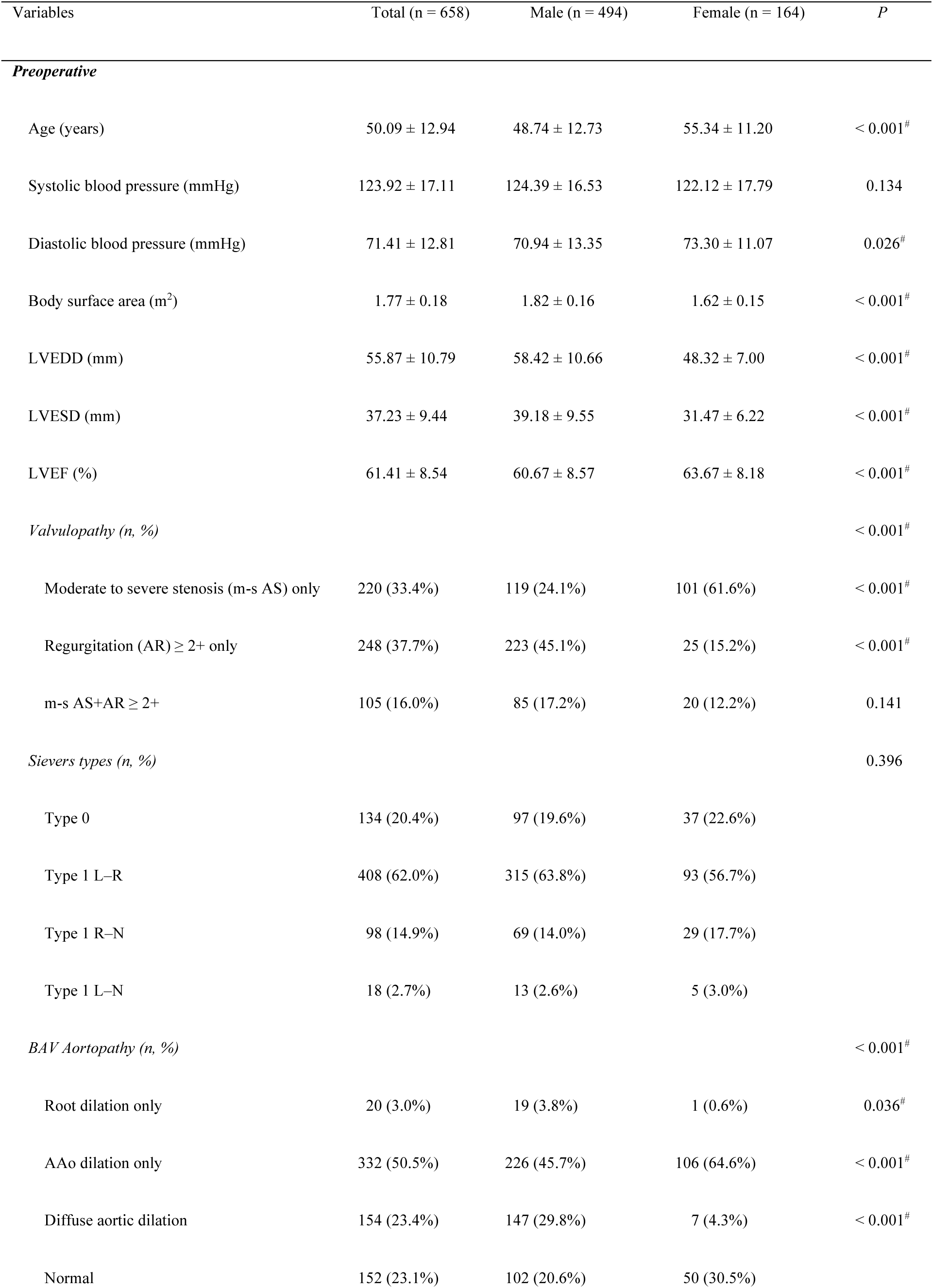
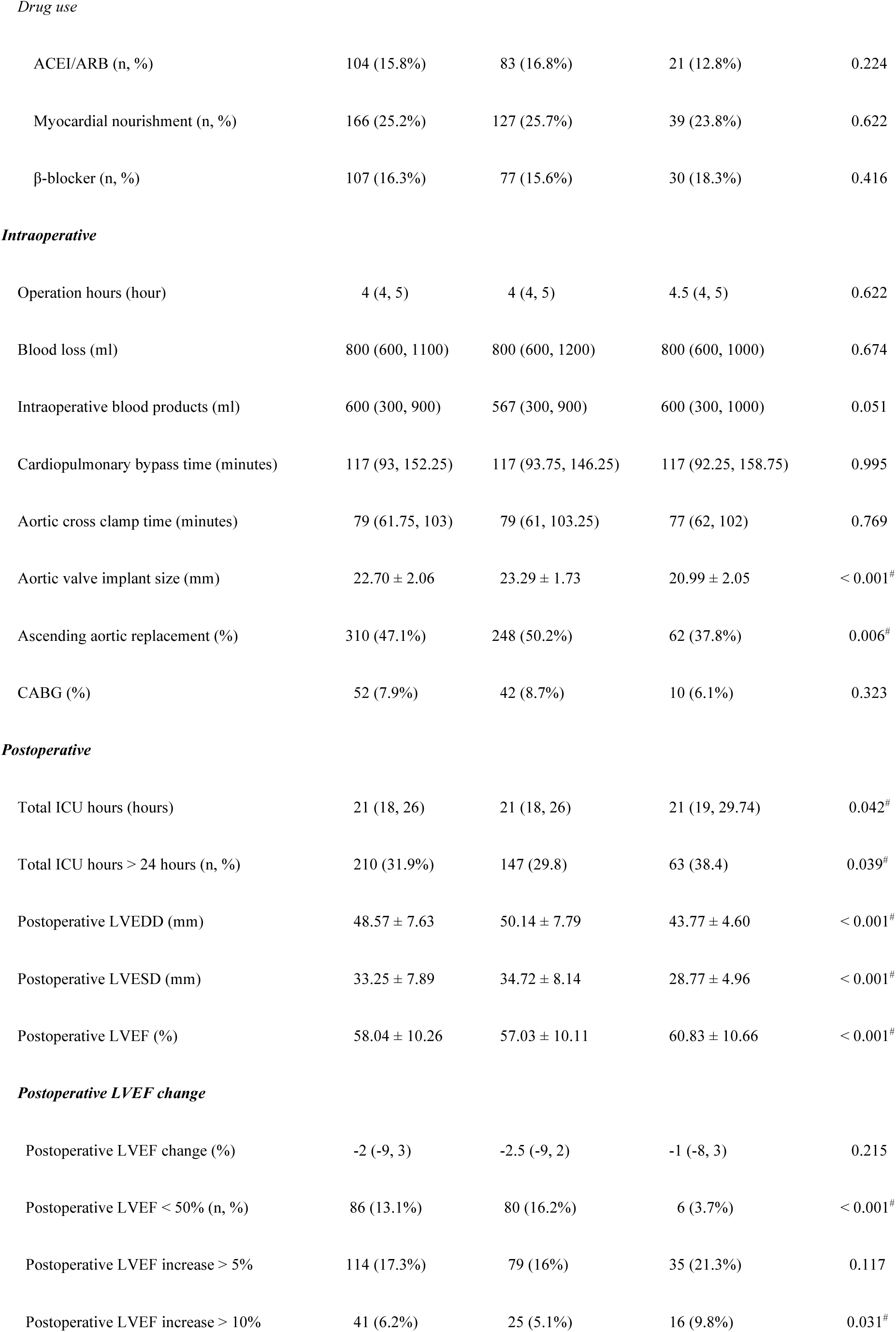
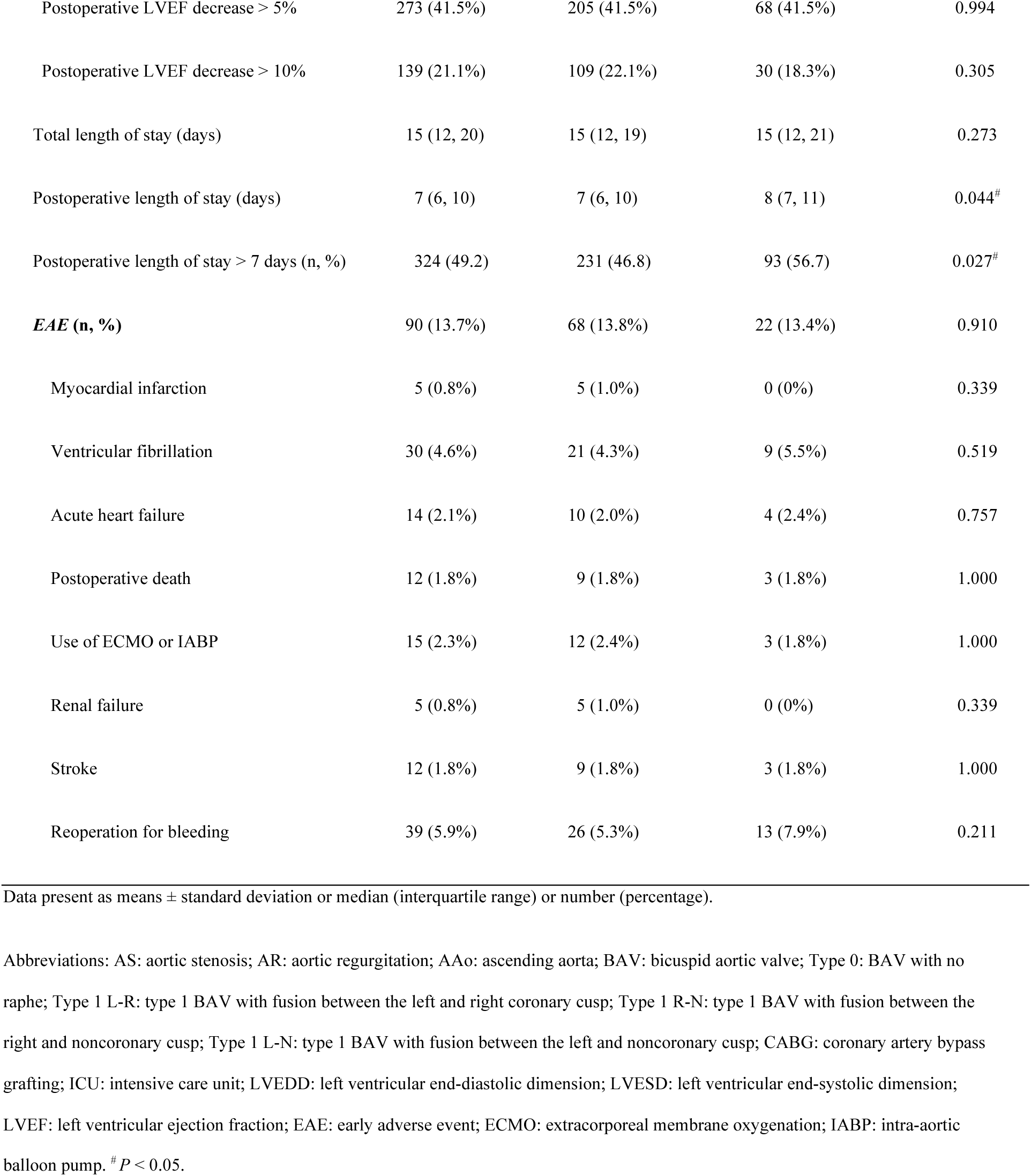
Summary of pre-, intra- and postoperative characteristics and outcomes of BAV patients underwent AVR

| Variables | Total (n = 658) | Male (n = 494) | Female (n = 164) | P |
| --- | --- | --- | --- | --- |
| *Preoperative* |  |  |  |  |
| Age (years) | 50.09 ± 12.94 | 48.74 ± 12.73 | 55.34 ± 11.20 | < 0.001<sup>#</sup> |
| Systolic blood pressure (mmHg) | 123.92 ± 17.11 | 124.39 ± 16.53 | 122.12 ± 17.79 | 0.134 |
| Diastolic blood pressure (mmHg) | 71.41 ± 12.81 | 70.94 ± 13.35 | 73.30 ± 11.07 | 0.026<sup>#</sup> |
| Body surface area (m<sup>2</sup>) | 1.77 ± 0.18 | 1.82 ± 0.16 | 1.62 ± 0.15 | < 0.001<sup>#</sup> |
| LVEDD (mm) | 55.87 ± 10.79 | 58.42 ± 10.66 | 48.32 ± 7.00 | < 0.001<sup>#</sup> |
| LVESD (mm) | 37.23 ± 9.44 | 39.18 ± 9.55 | 31.47 ± 6.22 | < 0.001<sup>#</sup> |
| LVEF (%) | 61.41 ± 8.54 | 60.67 ± 8.57 | 63.67 ± 8.18 | < 0.001<sup>#</sup> |
| *Valvulopathy (n, %)* |  |  |  | < 0.001<sup>#</sup> |
| Moderate to severe stenosis (m-s AS) only | 220 (33.4%) | 119 (24.1%) | 101 (61.6%) | < 0.001<sup>#</sup> |
| Regurgitation (AR) ≥ 2+ only | 248 (37.7%) | 223 (45.1%) | 25 (15.2%) | < 0.001<sup>#</sup> |
| m-s AS+AR ≥ 2+ | 105 (16.0%) | 85 (17.2%) | 20 (12.2%) | 0.141 |
| *Sievers types (n, %)* |  |  |  | 0.396 |
| Type 0 | 134 (20.4%) | 97 (19.6%) | 37 (22.6%) |  |
| Type 1 L–R | 408 (62.0%) | 315 (63.8%) | 93 (56.7%) |  |
| Type 1 R–N | 98 (14.9%) | 69 (14.0%) | 29 (17.7%) |  |
| Type 1 L–N | 18 (2.7%) | 13 (2.6%) | 5 (3.0%) |  |
| *BAV Aortopathy (n, %)* |  |  |  | < 0.001<sup>#</sup> |
| Root dilation only | 20 (3.0%) | 19 (3.8%) | 1 (0.6%) | 0.036<sup>#</sup> |
| AAo dilation only | 332 (50.5%) | 226 (45.7%) | 106 (64.6%) | < 0.001<sup>#</sup> |
| Diffuse aortic dilation | 154 (23.4%) | 147 (29.8%) | 7 (4.3%) | < 0.001<sup>#</sup> |
| Normal | 152 (23.1%) | 102 (20.6%) | 50 (30.5%) |  |
| *Drug use* |  |  |  |  |
| ACEI/ARB (n, %) | 104 (15.8%) | 83 (16.8%) | 21 (12.8%) | 0.224 |
| Myocardial nourishment (n, %) | 166 (25.2%) | 127 (25.7%) | 39 (23.8%) | 0.622 |
| β-blocker (n, %) | 107 (16.3%) | 77 (15.6%) | 30 (18.3%) | 0.416 |
| *Intraoperative* |  |  |  |  |
| Operation hours (hour) | 4 (4, 5) | 4 (4, 5) | 4.5 (4, 5) | 0.622 |
| Blood loss (ml) | 800 (600, 1100) | 800 (600, 1200) | 800 (600, 1000) | 0.674 |
| Intraoperative blood products (ml) | 600 (300, 900) | 567 (300, 900) | 600 (300, 1000) | 0.051 |
| Cardiopulmonary bypass time (minutes) | 117 (93, 152.25) | 117 (93.75, 146.25) | 117 (92.25, 158.75) | 0.995 |
| Aortic cross clamp time (minutes) | 79 (61.75, 103) | 79 (61, 103.25) | 77 (62, 102) | 0.769 |
| Aortic valve implant size (mm) | 22.70 ± 2.06 | 23.29 ± 1.73 | 20.99 ± 2.05 | < 0.001<sup>#</sup> |
| Ascending aortic replacement (%) | 310 (47.1%) | 248 (50.2%) | 62 (37.8%) | 0.006<sup>#</sup> |
| CABG (%) | 52 (7.9%) | 42 (8.7%) | 10 (6.1%) | 0.323 |
| *Postoperative* |  |  |  |  |
| Total ICU hours (hours) | 21 (18, 26) | 21 (18, 26) | 21 (19, 29.74) | 0.042<sup>#</sup> |
| Total ICU hours > 24 hours (n, %) | 210 (31.9%) | 147 (29.8) | 63 (38.4) | 0.039<sup>#</sup> |
| Postoperative LVEDD (mm) | 48.57 ± 7.63 | 50.14 ± 7.79 | 43.77 ± 4.60 | < 0.001<sup>#</sup> |
| Postoperative LVESD (mm) | 33.25 ± 7.89 | 34.72 ± 8.14 | 28.77 ± 4.96 | < 0.001<sup>#</sup> |
| Postoperative LVEF (%) | 58.04 ± 10.26 | 57.03 ± 10.11 | 60.83 ± 10.66 | < 0.001<sup>#</sup> |
| *Postoperative LVEF change* |  |  |  |  |
| Postoperative LVEF change (%) | -2 (-9, 3) | -2.5 (-9, 2) | -1 (-8, 3) | 0.215 |
| Postoperative LVEF < 50% (n, %) | 86 (13.1%) | 80 (16.2%) | 6 (3.7%) | < 0.001<sup>#</sup> |
| Postoperative LVEF increase > 5% | 114 (17.3%) | 79 (16%) | 35 (21.3%) | 0.117 |
| Postoperative LVEF increase > 10% | 41 (6.2%) | 25 (5.1%) | 16 (9.8%) | 0.031<sup>#</sup> |
| Postoperative LVEF decrease > 5% | 273 (41.5%) | 205 (41.5%) | 68 (41.5%) | 0.994 |
| Postoperative LVEF decrease > 10% | 139 (21.1%) | 109 (22.1%) | 30 (18.3%) | 0.305 |
| Total length of stay (days) | 15 (12, 20) | 15 (12, 19) | 15 (12, 21) | 0.273 |
| Postoperative length of stay (days) | 7 (6, 10) | 7 (6, 10) | 8 (7, 11) | 0.044<sup>#</sup> |
| Postoperative length of stay > 7 days (n, %) | 324 (49.2) | 231 (46.8) | 93 (56.7) | 0.027<sup>#</sup> |
| **EAE (n, %)** | **90 (13.7%)** | **68 (13.8%)** | **22 (13.4%)** | **0.910** |
| Myocardial infarction | 5 (0.8%) | 5 (1.0%) | 0 (0%) | 0.339 |
| Ventricular fibrillation | 30 (4.6%) | 21 (4.3%) | 9 (5.5%) | 0.519 |
| Acute heart failure | 14 (2.1%) | 10 (2.0%) | 4 (2.4%) | 0.757 |
| Postoperative death | 12 (1.8%) | 9 (1.8%) | 3 (1.8%) | 1.000 |
| Use of ECMO or IABP | 15 (2.3%) | 12 (2.4%) | 3 (1.8%) | 1.000 |
| Renal failure | 5 (0.8%) | 5 (1.0%) | 0 (0%) | 0.339 |
| Stroke | 12 (1.8%) | 9 (1.8%) | 3 (1.8%) | 1.000 |
| Reoperation for bleeding | 39 (5.9%) | 26 (5.3%) | 13 (7.9%) | 0.211 |

Data present as means ± standard deviation or median (interquartile range) or number (percentage).
Abbreviations: AS: aortic stenosis; AR: aortic regurgitation; AAo: ascending aorta; BAV: bicuspid aortic valve; Type 0: BAV with no raphe; Type 1 L-R: type 1 BAV with fusion between the left and right coronary cusp; Type 1 R-N: type 1 BAV with fusion between the right and noncoronary cusp; Type 1 L-N: type 1 BAV with fusion between the left and noncoronary cusp; CABG: coronary artery bypass grafting; ICU: intensive care unit; LVEDD: left ventricular end-diastolic dimension; LVESD: left ventricular end-systolic dimension; LVEF: left ventricular ejection fraction; EAE: early adverse event; ECMO: extracorporeal membrane oxygenation; IABP: intra-aortic balloon pump. <sup>#</sup> $P < 0.05$ .

Diffuse aortic dilation (OR = 2.899, *P* = 0.013), ascending aortic replacement (OR = 2.154, *P* = 0.016) and longer aortic cross clamp time (OR = 1.012, *P* < 0.001) were the risk factors for postoperative LVEF < 50%, while higher preoperative LVEF is the protective factor (OR = 0.873, *P* < 0.001) (table 6). Age (OR = 1.018, *P* = 0.015) and longer aortic cross clamp (ACC) time (OR = 1.015, *P* < 0.001) were the risk factors for total ICU hours > 24 hours. Higher SBP (OR = 0.990, *P* = 0.047) and accompanied ascending aortic replacement (OR = 0.552, *P* = 0.001) decrease the possibility of postoperative length of stay > 7 days, while m-s AS only (OR = 1.846, *P* = 0.027) and longer ACC time (OR = 1.005, *P* = 0.021) increased the risk of staying more days in the hospital.

**Table 6.** Factors related to post-OP EAE, postoperative LVEF < 50%, total ICU hours > 24 hours and Postoperative length of stay > 7 days

| Variables | Cox regression: EAE |  | Logistic regression: postoperative<br>LVEF<50% |  | Total ICU hours > 24 hours |  | Postoperative length of stay > 7 days |  |
| --- | --- | --- | --- | --- | --- | --- | --- | --- |
|  | Univariable | Multivariable | Univariable | Multivariable | Univariable | Multivariable | Univariable | Multivariable |
|  | analysis (HR, CI,<br>P) | analysis (HR, CI, P) | analysis (OR, CI,<br>P) | analysis (OR, CI, P) | analysis (OR, CI,<br>P) | analysis (OR, CI, P) | analysis (OR, CI,<br>P) | analysis (OR, CI, P) |
| Male | 1.542, 0.875– | 1.213, 0.742–1.982, | 3.358, 1.135– | 3.071, 1.096–8.606, | 0.650, 0.406– | 0.710, 0.479–1.053, | 0.754, 0.485– | 0.791, 0.533– |
|  | 2.717, 0.134 | 0.441 | 9.936, 0.029 | 0.033<sup>#</sup> | 1.043, 0.074 | 0.089 | 1.173, 0.210 | 1.173, 0.243 |
| Age | 1.028, 1.007– | 1.030, 1.011–1.050, | 1.006, 0.981– |  | 1.019, 1.004– | 1.018, 1.003–1.033, | 1.003, 0.989– |  |
|  | 1.049, 0.008 | 0.002<sup>#</sup> | 1.031, 0.648 |  | 1.036, 0.017 | 0.015<sup>#</sup> | 1.017, 0.645 |  |
| Body surface area | 0.439, 0.107– |  | 0.740, 0.132– |  | 2.119, 0.681– |  | 1.355, 0.484– |  |
|  | 1.799, 0.253 |  | 4.160, 0.740 |  | 6.597, 0.195 |  | 3.789, 0.563 |  |
| Systolic blood pressure | 0.995, 0.983– |  | 0.977, 0.955– | 0.976, 0.955–0.999, | 1.000, 0.989– |  | 0.990, 0.980– | 0.990, 0.981–1.000, |
|  | 1.009, 0.500 |  | 1.000, 0.046 | 0.037<sup>#</sup> | 1.010, -0.930 |  | 1.000, 0.404 | 0.047<sup>#</sup> |
| **Valvulopathy** |  |  |  |  |  |  |  |  |
| Moderate to severe<br>stenosis (m-s AS) only | 0.784, 0.405– |  | 0.785, 0.204– | 0.805, 0.213–3.052, | 0.904, 0.498– |  | 1.837, 1.052– | 1.846, 1.074–3.172, |
|  | 1.518, 0.471 |  | 3.014, 0.724 | 0.750 | 1.639, 0.739 |  | 3.207, 0.032 | 0.027<sup>#</sup> |
| Regurgitation (AR) ≥ 2+ only | 0.843, 0.416–<br>1.708, 0.635 |  | 3.031, 0.919–<br>9.990, 0.068 | 2.895, 0.903–9.287,<br>0.074 | 0.750, 0.417–<br>1.348, 0.337 |  | 1.060, 0.623–<br>1.802, 0.830 | 1.060, 0.628–1.790,<br>0.827 |
| m-s AS+AR≥ 2+ | 0.773, 0.359–<br>1.661, 0.509 |  | 1.335, 0.360–<br>4.950, 0.666 | 1.340, 0.368–4.877,<br>0.657 | 0.953, 0.496–<br>1.832, 0.885 |  | 0.977, 0.535–<br>1.786, 0.941 | 0.961, 0.530–1.745,<br>0.897 |
| ***BAV Aortopathy*** |  |  |  |  |  |  |  |  |
| Root dilation only | 0.626, 0.080–<br>4.887, 0.655 |  | 1.526, 0.338–<br>6.890, 0.583 | 1.453, 0.326–6.470,<br>0.624 | 1.509, 0.527–<br>4.326, 0.444 |  | 1.260, 0.477–<br>3.329, 0.641 |  |
| AAo dilation only | 1.480, 0.799–<br>2.740, 0.213 |  | 1.708, 0.769–<br>3.793, 0.188 | 1.751, 0.808–3.794,<br>0.156 | 0.834, 0.513–<br>1.356, 0.464 |  | 0.990, 0.634–<br>1.545, 0.963 |  |
| Diffuse aortic dilation | 0.867, 0.382–<br>1.968, 0.732 |  | 2.843, 1.179–<br>6.860, 0.020 | 2.899, 1.257–6.683,<br>0.013<sup>#</sup> | 0.620, 0.332–<br>1.159, 0.134 |  | 0.978, 0.560–<br>1.707, 0.938 |  |
| Preoperative LVEF | 1.003, 0.978–<br>1.028 0.828 |  | 0.873, 0.844–<br>0.903, 0.000 | 0.873, 0.845–0.902,<br>0.000<sup>#</sup> | 0.986, 0.966–<br>1.007, 0.199 |  | 0.999, 0.979–<br>1.018, 0.892 |  |
| Ascending aortic replacement | 0.944, 0.587–<br>1.518, 0.813 |  | 2.176, 1.159–<br>4.088, 0.016 | 2.154, 1.153–4.027,<br>0.016<sup>#</sup> | 0.857, 0.572–<br>1.282, 0.452 |  | 0.552, 0.381–<br>0.800, 0.002 | 0.552, 0.395–0.772,<br>0.001<sup>#</sup> |
| Aortic cross clamp time | 1.012, 1.008–<br>1.016, 0.000 | 1.012, 1.008–1.015,<br>0.000<sup>#</sup> | 1.012, 1.006–<br>1.019, 0.000 | 1.012, 1.006–1.019,<br>0.000<sup>#</sup> | 1.015, 1.010–<br>1.021, 0.000 | 1.015, 1.011–1.020,<br>0.000<sup>#</sup> | 1.005, 1.001–<br>1.010, 0.023 | 1.005, 1.001–1.010,<br>0.021<sup>#</sup> |

Abbreviations: AS: aortic stenosis; AR: aortic regurgitation; AAo: ascending aorta; LVEF: left ventricular ejection fraction; EAE: early adverse event. <sup>#</sup> $P < 0.05$ .

Furthermore, we analyzed the factors for postoperative LVEF improvements or deterioration by LVEF changes of 5% or 10% (table 7). Higher SBP (OR = 0.984, *P* = 0.017) and preoperative LVEF (OR = 0.880, *P* < 0.001), and accompanied ascending aortic replacement (OR = 0.604, *P* = 0.037) might hinder the patients from LVEF improvement by > 5%, while patients with m-s AS only (OR = 2.559, *P* = 0.018) were prone to have postoperative LVEF increase > 5%. Higher preoperative LVEF (OR = 0.855, *P* < 0.001) also decrease the chance of LVEF improvement by > 10%. Higher preoperative LVEF and AR ≥ 2+ only were risk factors for postoperative LVEF decrease > 5% (preoperative LVEF, OR = 1.120, *P* < 0.001; AR ≥ 2+ only, OR = 2.486, *P* = 0.001) or decrease > 10% (preoperative LVEF, OR = 1.110, *P* < 0.001; AR ≥ 2+ only, OR = 3.172, *P* = 0.001), while m-s AS only (OR = 0.437, *P* = 0.030) was the protective factor for postoperative LVEF decrease > 10%. (table 7).

**Table 7.**
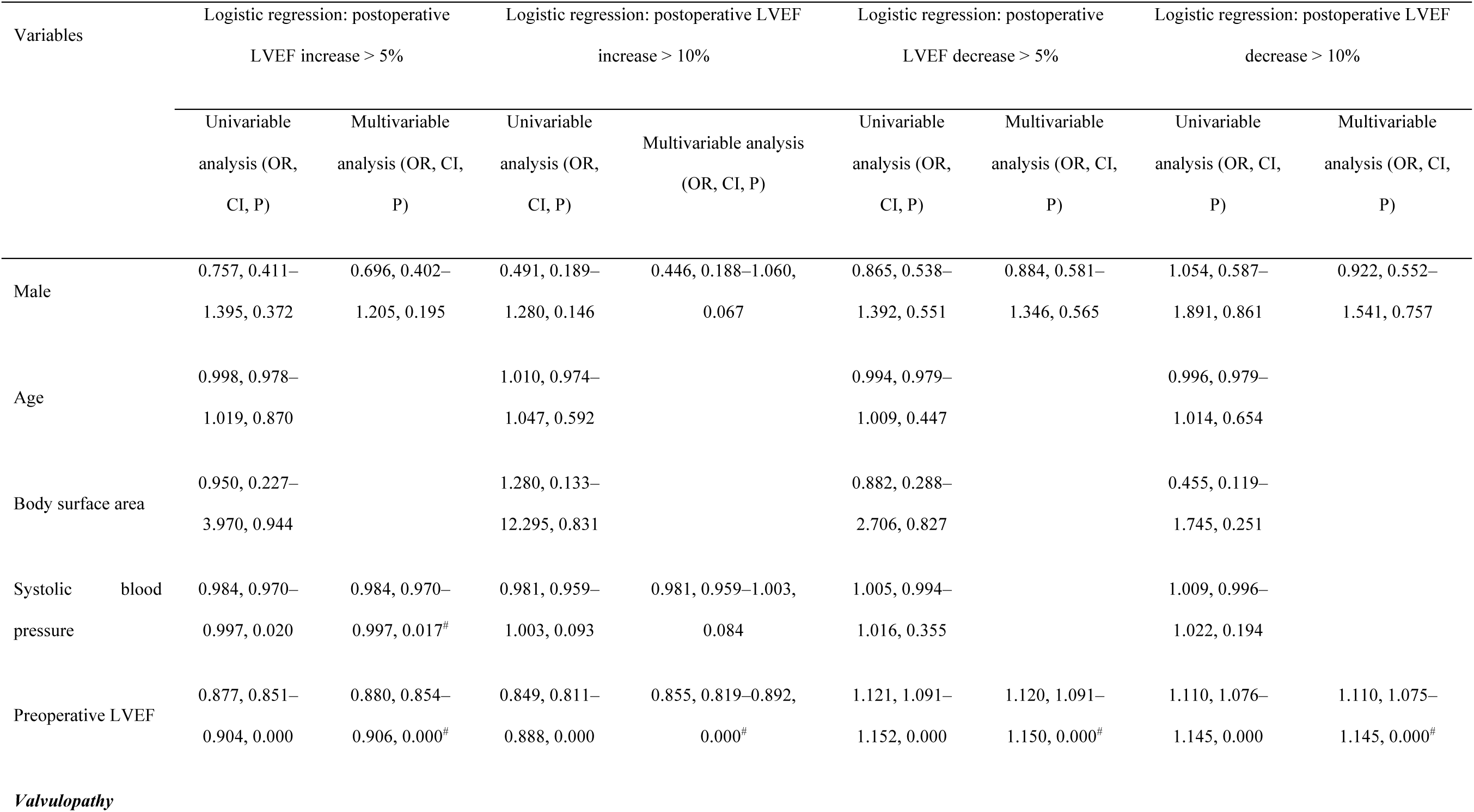
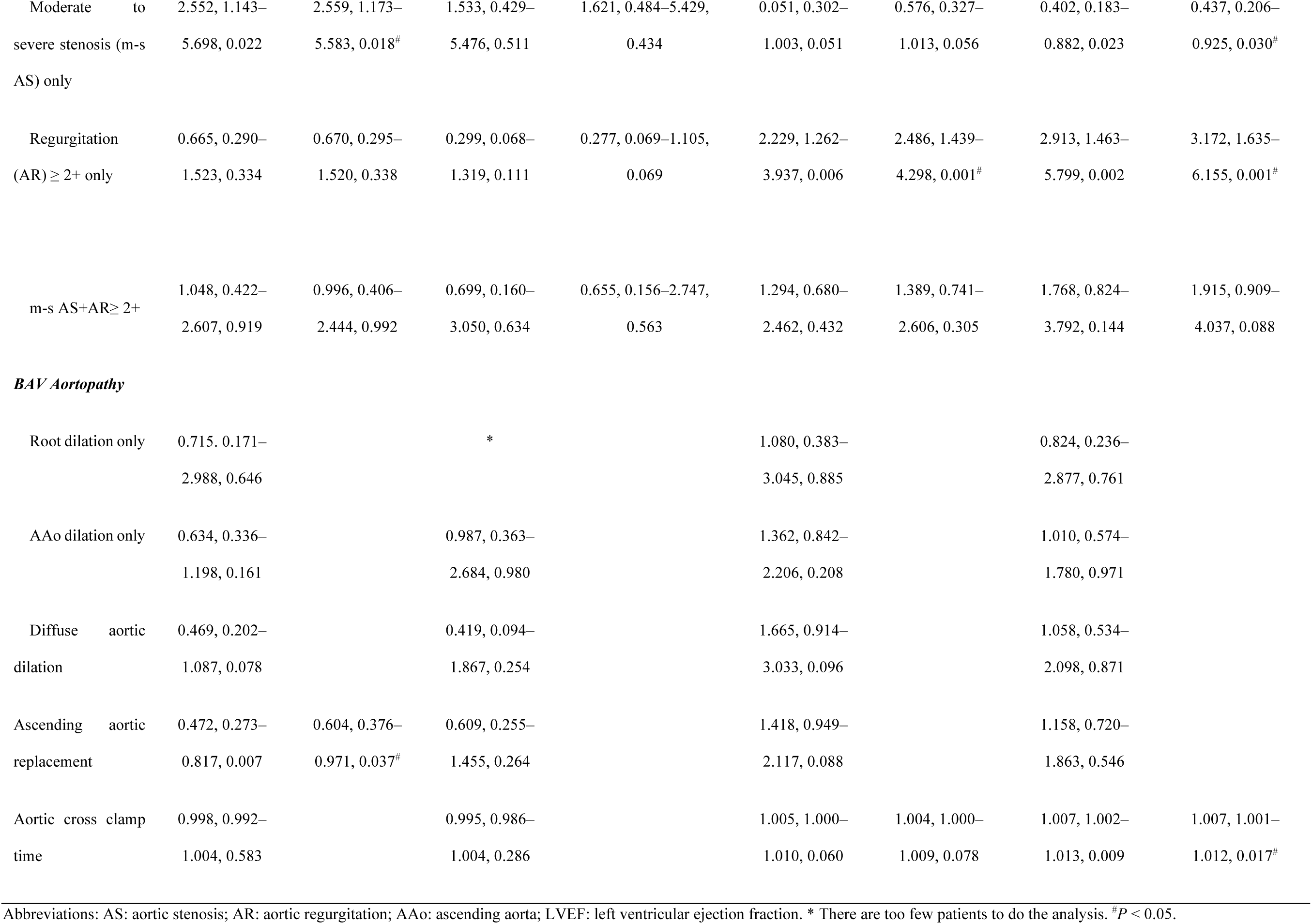
Factors related to postoperative LVEF changes

| Variables | Logistic regression: postoperative<br>LVEF increase > 5% |  | Logistic regression: postoperative LVEF<br>increase > 10% |  | Logistic regression: postoperative<br>LVEF decrease > 5% |  | Logistic regression: postoperative LVEF<br>decrease > 10% |  |
| --- | --- | --- | --- | --- | --- | --- | --- | --- |
|  | Univariable | Multivariable | Univariable | Multivariable analysis<br>(OR, CI, P) | Univariable | Multivariable | Univariable | Multivariable |
|  | analysis (OR,<br>CI, P) | analysis (OR, CI,<br>P) | analysis (OR,<br>CI, P) |  | analysis (OR,<br>CI, P) | analysis (OR, CI,<br>P) | analysis (OR, CI,<br>P) | analysis (OR, CI,<br>P) |
| Male | 0.757, 0.411– | 0.696, 0.402– | 0.491, 0.189– | 0.446, 0.188–1.060, | 0.865, 0.538– | 0.884, 0.581– | 1.054, 0.587– | 0.922, 0.552– |
|  | 1.395, 0.372 | 1.205, 0.195 | 1.280, 0.146 | 0.067 | 1.392, 0.551 | 1.346, 0.565 | 1.891, 0.861 | 1.541, 0.757 |
| Age | 0.998, 0.978– |  | 1.010, 0.974– |  | 0.994, 0.979– |  | 0.996, 0.979– |  |
|  | 1.019, 0.870 |  | 1.047, 0.592 |  | 1.009, 0.447 |  | 1.014, 0.654 |  |
| Body surface area | 0.950, 0.227– |  | 1.280, 0.133– |  | 0.882, 0.288– |  | 0.455, 0.119– |  |
|  | 3.970, 0.944 |  | 12.295, 0.831 |  | 2.706, 0.827 |  | 1.745, 0.251 |  |
| Systolic blood pressure | 0.984, 0.970– | 0.984, 0.970– | 0.981, 0.959– | 0.981, 0.959–1.003, | 1.005, 0.994– |  | 1.009, 0.996– |  |
|  | 0.997, 0.020 | 0.997, 0.017<sup>#</sup> | 1.003, 0.093 | 0.084 | 1.016, 0.355 |  | 1.022, 0.194 |  |
| Preoperative LVEF | 0.877, 0.851– | 0.880, 0.854– | 0.849, 0.811– | 0.855, 0.819–0.892, | 1.121, 1.091– | 1.120, 1.091– | 1.110, 1.076– | 1.110, 1.075– |
|  | 0.904, 0.000 | 0.906, 0.000<sup>#</sup> | 0.888, 0.000 | 0.000<sup>#</sup> | 1.152, 0.000 | 1.150, 0.000<sup>#</sup> | 1.145, 0.000 | 1.145, 0.000<sup>#</sup> |
| *Valvulopathy* |  |  |  |  |  |  |  |  |
| Moderate to severe stenosis (m-s AS) only | 2.552, 1.143–<br>5.698, 0.022 | 2.559, 1.173–<br>5.583, 0.018<sup>#</sup> | 1.533, 0.429–<br>5.476, 0.511 | 1.621, 0.484–5.429,<br>0.434 | 0.051, 0.302–<br>1.003, 0.051 | 0.576, 0.327–<br>1.013, 0.056 | 0.402, 0.183–<br>0.882, 0.023 | 0.437, 0.206–<br>0.925, 0.030<sup>#</sup> |
| Regurgitation (AR) ≥ 2+ only | 0.665, 0.290–<br>1.523, 0.334 | 0.670, 0.295–<br>1.520, 0.338 | 0.299, 0.068–<br>1.319, 0.111 | 0.277, 0.069–1.105,<br>0.069 | 2.229, 1.262–<br>3.937, 0.006 | 2.486, 1.439–<br>4.298, 0.001<sup>#</sup> | 2.913, 1.463–<br>5.799, 0.002 | 3.172, 1.635–<br>6.155, 0.001<sup>#</sup> |
| m-s AS+AR ≥ 2+ | 1.048, 0.422–<br>2.607, 0.919 | 0.996, 0.406–<br>2.444, 0.992 | 0.699, 0.160–<br>3.050, 0.634 | 0.655, 0.156–2.747,<br>0.563 | 1.294, 0.680–<br>2.462, 0.432 | 1.389, 0.741–<br>2.606, 0.305 | 1.768, 0.824–<br>3.792, 0.144 | 1.915, 0.909–<br>4.037, 0.088 |
| **BAV Aortopathy** |  |  |  |  |  |  |  |  |
| Root dilation only | 0.715, 0.171–<br>2.988, 0.646 |  | * |  | 1.080, 0.383–<br>3.045, 0.885 |  | 0.824, 0.236–<br>2.877, 0.761 |  |
| AAo dilation only | 0.634, 0.336–<br>1.198, 0.161 |  | 0.987, 0.363–<br>2.684, 0.980 |  | 1.362, 0.842–<br>2.206, 0.208 |  | 1.010, 0.574–<br>1.780, 0.971 |  |
| Diffuse aortic dilation | 0.469, 0.202–<br>1.087, 0.078 |  | 0.419, 0.094–<br>1.867, 0.254 |  | 1.665, 0.914–<br>3.033, 0.096 |  | 1.058, 0.534–<br>2.098, 0.871 |  |
| Ascending aortic replacement | 0.472, 0.273–<br>0.817, 0.007 | 0.604, 0.376–<br>0.971, 0.037<sup>#</sup> | 0.609, 0.255–<br>1.455, 0.264 |  | 1.418, 0.949–<br>2.117, 0.088 |  | 1.158, 0.720–<br>1.863, 0.546 |  |
| Aortic cross clamp time | 0.998, 0.992–<br>1.004, 0.583 |  | 0.995, 0.986–<br>1.004, 0.286 |  | 1.005, 1.000–<br>1.010, 0.060 | 1.004, 1.000–<br>1.009, 0.078 | 1.007, 1.002–<br>1.013, 0.009 | 1.007, 1.001–<br>1.012, 0.017<sup>#</sup> |

Abbreviations: AS: aortic stenosis; AR: aortic regurgitation; AAo: ascending aorta; LVEF: left ventricular ejection fraction. \* There are too few patients to do the analysis. <sup>#</sup> $P < 0.05$ .

## Discussion

In this study, we performed a comprehensive evaluation of the relationships among demographic and clinical characteristics, BAV Sievers types, valvulopathy and aortopathy, and the risk/protective factors for outcomes after aortic valve replacement (AVR) in BAV patients, with our large retrospective cohort of BAV patients. Our study unveiled multiple previously unrecognized interwoven interactions between some demographic characteristics, BAV Sievers types, and operational outcomes.

### Demographic characteristics’ effects on BAV associated valvulopathy and aortopathy

Though it is well known that BAV is a male predominant entity^4, 23^, as also shown by our study, whether there are sex differences in BAV Sievers types, valvulopathy, aortopathy and operational outcomes has been controversial. Our study demonstrated that type 1 L–R was the most prevalent configuration without sex difference in Sievers types, agreeing with the reports by Kong et al.^4^ and by Lee et al^5^. On the contrary, Roman et al. reported that among 424 unoperated BAV patients, males had more frequently type 1 L - R than females (81.5% vs. 69.0%, *P* = 0.03), whereas type 1 L - N was more common in females (18.5% vs. 31.0%, *P* = 0.03)^3^. In addition, our cohort showed that male BAV patients were younger, with larger BSA, LVEDD and LVESD, and lower LVEF than females and had more need for AVR or AVR+AAR, suggesting faster development and worse outcomes of aortopathy and valvulopathy in males.

### Influences of Demographic and clinical characteristics, and BAV Sievers types on aortopathy and valvulopathy in BAV patients

As BAV is associated with adverse aortopathy and valvulopathy which have significant impact on the health of these patients, it is necessary to test if the demographic and clinical characteristics, like age, sex, BSA and blood pressure, and Sievers types were independent predictors for BAV valvulopathy, aortopathy. One advantage of our study is that the large number of patients and comprehensive dataset enabled us to analyze risk/protective factors of aortopathy and valvulopathy in subpopulations such as male patients or patients with specific comorbidities.

Sievers types and sex may affect aortopathy. Sievers et al^24^ showed that AAo dilation was observed more often in BAV type 1 L-R and type 0 than in other Sievers types. Aortic root dilation was more often in type 0 and type 1 R-N, while diffuse aortic dilation was more frequent in type 1 L-R. Our results also showed patients with type 1 L-R had the highest proportion of diffuse aortic dilation, but we found that type 1 R-N but not type 0 was more likely to have AAo dilation only, which was different from Sievers’ result. Our discovery that the frequency of m-s AS only was similar in different Sievers types was also different from the results reported by Sievers et al^24^. These differences could be due to the fact that different BAV fusion pattern may lead to the hemodynamic changes in aorta in distinct ways, resulting in different types of aortopathy^25, 26^. In terms of the effects of sex, Kong et al. pointed out that males showed more frequent aortic SOV dilation compared with females^4^. Andrei et al. reported males with BAV had larger aortic dimension but did not mention which segment of the aorta was dilated^6^. In our study, we examined aortic dilation both at the root and ascending segments. The diameter of SOV, AAo, BSA indexed SOV were larger in males, but normalized AAo by BSA was larger in females. Aortic root dilation only or diffuse aortic dilation was more frequently found in males while females tended to have more AAo dilation. In addition, in our whole cohort, higher BSA, age, m-s AS+AR≽ 2+ were the risk factors for all or some types of aortopathy but m-s AS only was a protective factor for diffuse aortic dilation. Male shared similar prediction factors of aortopathy with the whole cohort. But in females, only age could be determined as an independent predictor for AAo dilation only.

Valvulopathy was in 73.4% of BAV patients in our study cohort (m-s AS and/or AR ≥ 2+). We found that male BAV patients were present more often with m-s AR only at a young age, while female BAV patients had more frequent AS, consistent with the result of a Krepps’s stduy^3^ and Kong’s study^4^. Kong’s study only examined BAV Sievers types and sex on types of valvulopathy and concluded that only sex was related. Our data presented a much more complex situation: for the whole cohort, age is the risk factor for m-s AS only but higher BSA and diffuse aortic dilation were the protective factors. Higher SBP and male would increase the risk of AR ≥ 2+ only. For m-s AS +AR ≥ 2+, AAo dilation, diffuse aortic dilation and male were the risk factor while higher BSA would protect the patients from m-s AS +AR ≥ 2+. The risk or protect factors of valvulopathy in male were similar as for whole cohort. But in females, the only identified risk factor for m-s AS only was age, while age and BSA were independent determinants for m-s AS +AR ≥ 2+.

### Biological mechanisms _ for aortopathy and valvulopathy associated with BAV

There were two main hypotheses to explain aortic dilation: hemodynamic theory and gene theory^27-29^. In AS patient, the aortic dilation could be induced by valve related flow turbulence (hemodynamic theory), while AR could be the results of aortic root dilation caused by related gene mutations (gene theory), as suggested by Sievers et al^24^. In our study, we showed sex difference in aortopathy though not the same as previously reported, consistent with the “X chromosome protection theory:^30-32^ Further investigation is needed to explain these sex differences in BAV aortopathy.

The mechanisms for BAV valvulopathy are not clear. Gene mutations or expression changes causes BAV Sievers types, which may be directly associated with different subtypes of valvulopathy^33^. In our study, male sex is also related to AR ≥ 2+ or m-s AS+ AR ≥ 2+, suggesting some genetic influence on the development of specific types of valvulopathy.

### Who can obtain more benefits or are exposed to more risks to surgical interventions in BAV patients?

Our study has suggested that male BAV patients advance to more severe (e.g., lower LVEF) conditions that even requires surgical interventions at younger ages. So, it seems that male BAV patients should benefit more from surgical interventions. However, we found no significant difference in ACC time and EAE between sex though females needed longer total ICU hours and postoperative length of stay. In contrast, Andrei and colleagues implied that females had higher risk for in-hospital mortality and longer ICU median hours of stay, longer ACC times and higher preoperative LVEF^6^. Another study analyzed 3 cohort (community cohort, tertiary clinical referral cohort, and surgical referral cohort), and found that in surgical referral cohort, females exhibited a higher relative risk of death^10^. We found that age and longer ACC time were risk factors for EAE and total ICU hours > 24 hours.

We further evaluated the benefits and risks of surgical interventions by examining the more subtle but critical parameters, postoperative LVEF < 50% and LVEF change by operation and total post-operation stay. Postoperative LVEF < 50% could be a predictor for 5-year survival for AVR in BAV patients^22^. Male, lower preoperative LVEF, accompanied ascending aortic replacement and longer ACC time were risk factor of postoperative LVEF < 50%. However, the lower preoperative LVEF was also the predictor for postoperative LVEF increase. AS only patients were prone to have LVEF increase after surgery which agrees the AVR in BAV/TAV patients^11, 34^ probably due to the decrease of afterload after the removal of stenosed AV. AR patients were prone to have postoperative LVEF decrease as reported by Disha et al.^11^, which could reflect no need of high LVEF after regurgitation correction. For longer postoperative stay (>7 days), AS, age and longer ACC time were risk factors, but accompanied ascending aortic replacement was shown to be a protective factor.

## Study limitations

The principal limitation of this study was its single center based retrospective study, and the short follow-up time after surgeries.

This study is based on a single center though our patient number is large. Also, we only analyzed hospitalized patients, and among them, surgical inpatients occupy a large proportion. This might lead to a selection bias. Besides, the patients enrolled in this study were all Chinese (Asians), and it was known that there were some anatomical and genetic differences between American, European and Asian patients.^35^ By far, only few studies were available for the characterization of BAV patients of Chinese or Asians ^35-37^.We have already registered a multicenter study on BAV (registration number: ChiCTR-RRC-17013678) which will enable us to further test our findings and to carry out the long-term follow-up in the future.

There were several data analyses that cannot be done because of the limited number of patients in subgroups such as some Sievers types or a subtype of aortopathy in females, AAo dilation only in females, m-s AS +AR≥ 2+ and root dilation only in whole cohort and males.

## Conclusion

Our study provided the evidence for comprehensive relationships between demographic and clinical characteristics, BAV Sievers types, valvulopathy and aortopathy, and the possible risk factors for adverse outcomes after AVR in BAV patients. Sex, SBP, age, Sievers types, interactions between aortopathy and valvulopathy differently impact on aortopathy, valvulopathy and the short outcomes of AVR.

Author list:

Yijia Li

Department of Echocardiography, Beijing Anzhen Hospital, Capital Medical University, and Beijing Institute of Heart Lung and Blood Vessel Diseases

Department of Physiology & Cardiovascular Research Center, Temple University School of Medicine

phone: +1 2677522688

Qiong Zhao:

Qiong Zhao: Institute: Inova Heart and Vascular Institute, Inova Fairfax Hospital, Cardiac Diagnostic, 1st Floor, 3300 Gallow’s Road, Falls Church, VA 22042, USA;

Phone: 5714722900

Yue Qi:

Department of Epidemiology, Beijing Anzhen Hospital, Capital Medical University, and Beijing Institute of Heart Lung and Blood Vessel Diseases

phone: +86 13811106169

Yichen Qu

Department of Echocardiography, Beijing Anzhen Hospital, Capital Medical University, and Beijing Institute of Heart Lung and Blood Vessel Diseases

phone: +86 15959146872

Akshay Kumar

Address: Department of Cardiothoracic Surgery, Medanta Hospital, Gurugram, India, Medanta Hospital, Islampur Colony, Sector 38, Gurugram, India 122001

phone: 843-870-3792

Ya Yang:

Department of Echocardiography, Beijing Anzhen Hospital, Capital Medical University, and Beijing Institute of Heart Lung and Blood Vessel Diseases

E-mail: echoyangya6666@ 163.com

phone: +86 18911662958

Xiongwen Chen:

Department of Physiology & Cardiovascular Research Center, Temple University School of Medicine

Phone: +1 215-707-3542

## Data Availability

All data referred to in the manuscript are available upon request.

## Acknowledgment

Dr. Li had full access to all of the data in the study and takes responsibility for the integrity of the data and the accuracy of the data analysis. Dr. Qi and Qu carried out the statistical analysis; Dr. Zhao helped with study concept and design. Dr. Chen and Yang proposed the study directions, suggested critical analyses, and performed critical revision.

---

**Supplementary tables:**

**Table 1.**
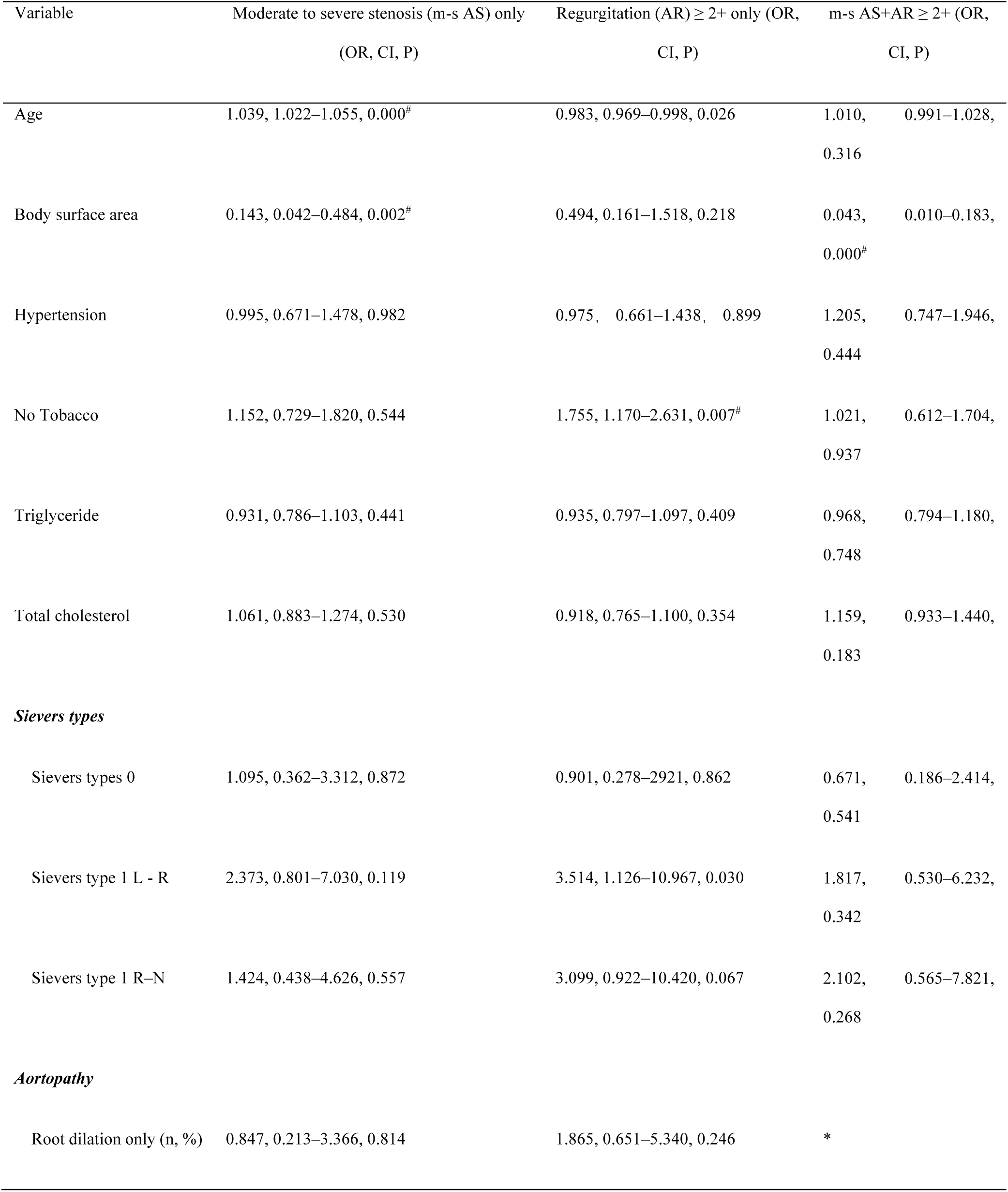
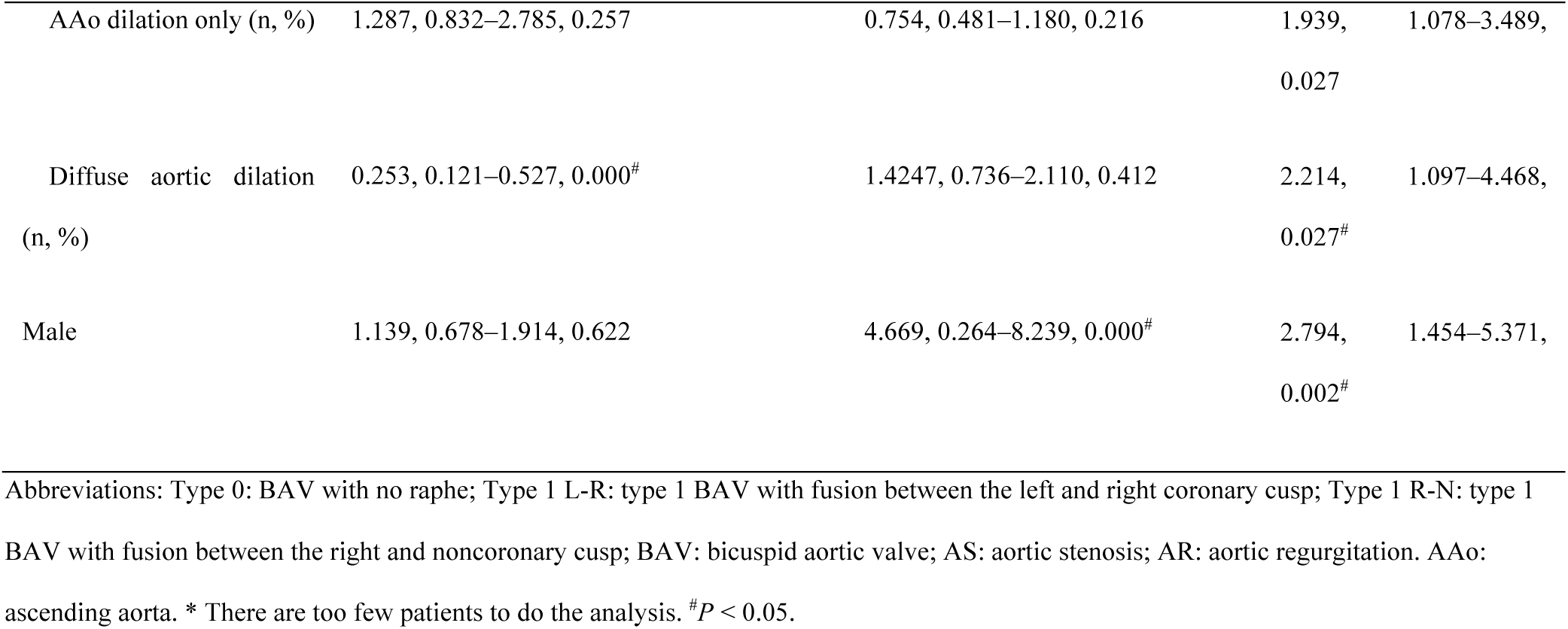
Risk factors for BAV Valvulopathy (use hypertension instead of systolic blood pressure)

**Table 2.**
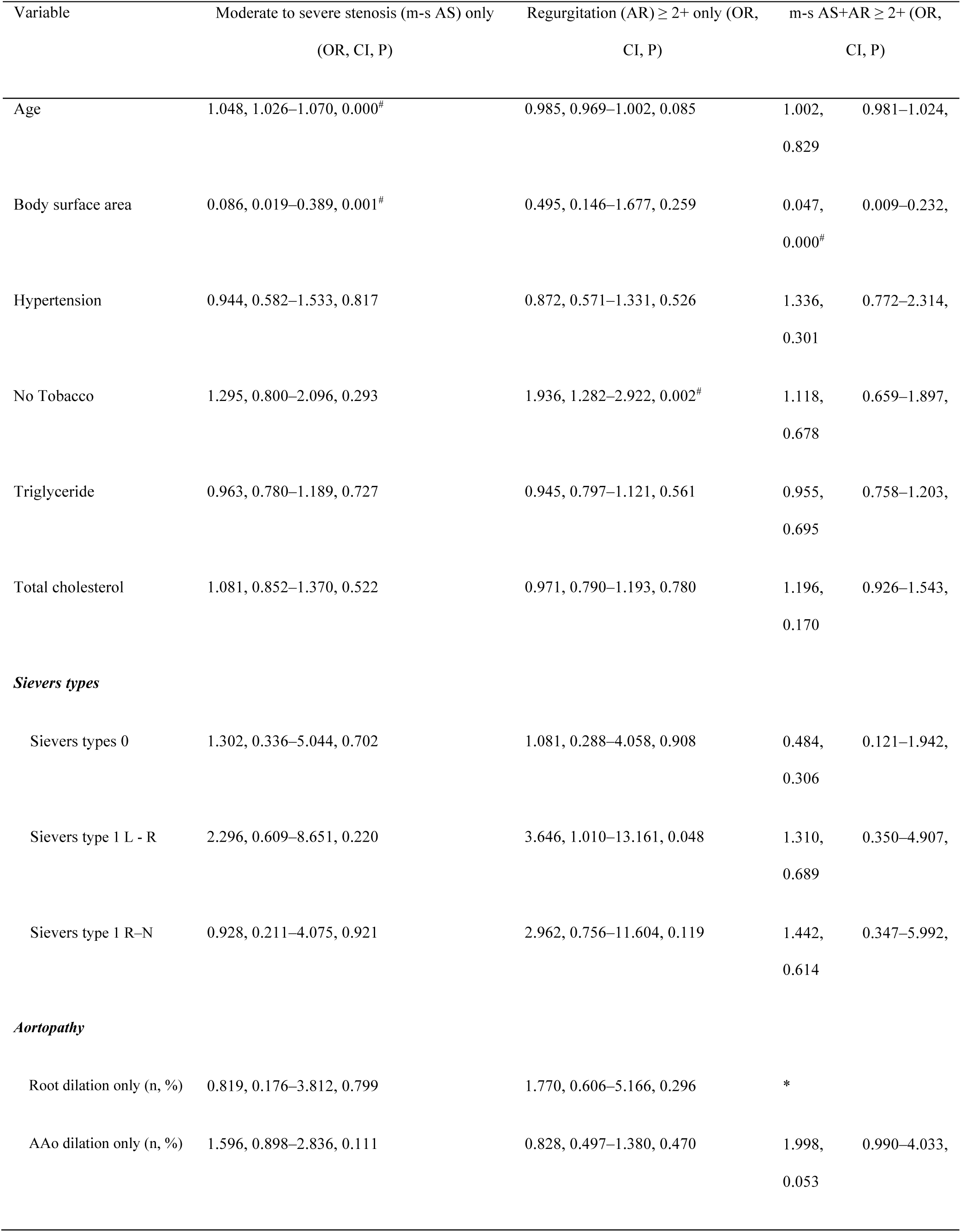
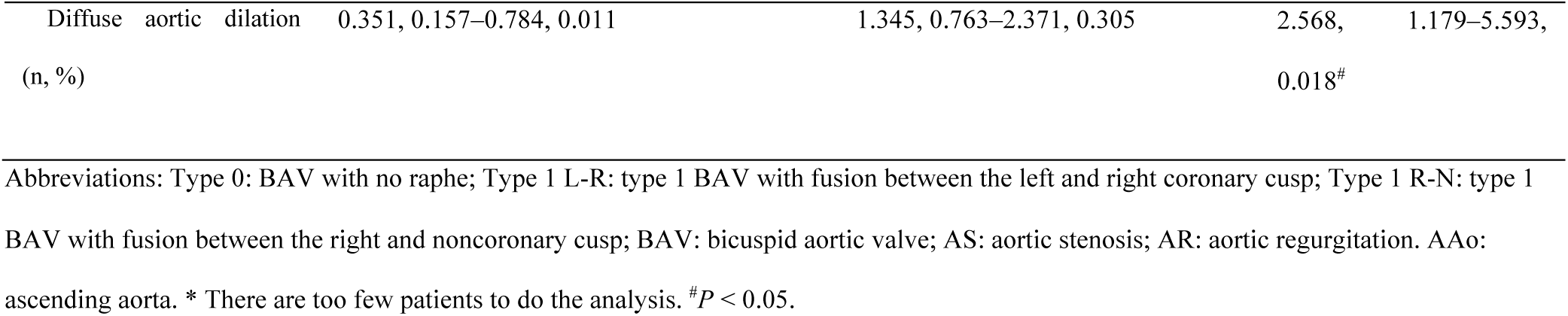
Risk factors for BAV Valvulopathy in male (use hypertension instead of systolic 1 blood pressure)

